# Apparent RSV–COVID interference is not robust to adjustment for shared testing propensity

**DOI:** 10.64898/2025.12.30.25343230

**Authors:** Joshua Steier

## Abstract

**Background:** Viral interference, in which infection by one pathogen reduces susceptibility to another at the population level, may shape respiratory virus dynamics. Inference from surveillance data is complicated by time-varying testing behavior that can induce correlated detection patterns without any biological interaction.

**Methods:** I developed a two-pathogen renewal model augmented with a ratio penalty that constrains interference estimates to be consistent with observed log-odds ratios of pathogen positivity. The penalty treats other-pathogen positives as implicit controls for shared testing propensity, adapting test-negative design logic to aggregate surveillance. I applied the model to US NAAT surveillance data reported to NREVSS (RSV and COVID-19; October 2020 to February 2026), validated parameter recovery in synthetic experiments, and quantified uncertainty via block bootstrap. I note at the outset that the method is conservative by design: synthetic experiments confirm a bias toward null interference estimates, so near-zero findings should not be read as proof that interference is absent.

**Results:** Without the ratio penalty, estimated interference was |*θ*|_sum_ = 0.0082 for RSV →COVID. With the penalty, this decreased to 0.0016 (80% reduction). Bootstrap 95% intervals included zero for all direction ×lag combinations. Synthetic validation confirmed high specificity at *θ* = 0 but revealed that the method cannot recover moderate interference (*θ ≤* 0.05), because virus-specific transmissibility deviations absorb the interference signal during Stage 1 estimation. A diagnostic decomposition showed that the ratio penalty term amplifies this bias-to-null: at *θ* = 0.01 in real data, the ratio penalty contributes a *−*314,000 log-joint penalty, roughly 130 times the multinomial penalty alone. Two-stage estimation was justified empirically; joint MAP estimation failed to converge across all tested configurations.

**Conclusions:** The ratio penalty functions as a conservative diagnostic screen with high specificity but limited sensitivity. When applied to RSV–COVID surveillance, it substantially reduces interference point estimates, with confidence intervals spanning zero. These results indicate that apparent interference signals in these data are not robust to this particular adjustment, but the method’s known conservative bias means biological interference cannot be excluded. The approach is best understood as a sensitivity analysis rather than a definitive test.

**Author Summary:** When one respiratory virus circulates widely, it may temporarily suppress transmission of others, a phenomenon called viral interference. Detecting interference from disease surveillance data is difficult because testing behavior changes over time: when any respiratory illness surges, more people seek tests, potentially creating correlated patterns that mimic biological interaction.

I developed a statistical method to probe this confounding. Borrowing logic from vaccine studies, the method penalizes the model when its predictions diverge from the observed ratio of positive tests across pathogens. The idea is that this ratio should be stable if testing propensity fluctuates but affects all pathogens similarly.

Applied to five years of US surveillance data for RSV and COVID-19, this penalty reduced apparent interference by 80%, with statistical uncertainty intervals including zero. Crucially, the method is intentionally conservative: simulation experiments show it also diminishes real interference signals, because transmissibility parameters absorb the interference effect before it can be estimated. My near-zero estimates therefore do not prove interference is absent; rather, they indicate that apparent signals in these data are not robust to this particular adjustment for testing composition.

This work highlights that surveillance-based interference estimates may be sensitive to testing artifacts and provides one approach for assessing this sensitivity.

## 1 Introduction

Respiratory viruses cause substantial morbidity worldwide [1, 2]. The emergence of SARS-CoV-2 raised questions about how multiple respiratory pathogens interact when co-circulating [5, 6].

Viral interference refers to the phenomenon whereby circulation of one pathogen at the population level is associated with reduced transmission of another. Proposed biological mechanisms operate at the individual level, including innate immune activation through interferon responses that confer temporary, non-specific protection against heterologous infection [7, 8]. Whether and how these individual-level mechanisms aggregate to produce detectable population-level effects depends on the prevalence of recently infected individuals, the duration and breadth of cross-protection, and the structure of contact patterns [5, 9]. In this paper, I use “interference” to denote population-level suppression of transmission, acknowledging that this may or may not reflect individual-level biological mechanisms.

If interference effects are substantial, they could influence epidemic timing and complicate pandemic preparedness [9, 10]. Studies have reported evidence for respiratory virus interference using clinical and surveillance data [5, 11, 12]. However, findings are heterogeneous: Zhang et al. found RSV–COVID interference to be weak or absent [13], and Waterlow et al. demonstrated fundamental identifiability limitations when estimating interaction strength from surveillance alone [17]. Several recent studies have advanced multi-pathogen modeling approaches for this problem [24–27]. This variability across studies motivates closer examination of the methods used for inference.

A fundamental challenge is distinguishing biological interactions from measurement artifacts. Respiratory virus surveillance relies on diagnostic testing, and testing behavior varies over time in ways that correlate across pathogens [14]. During surges, symptomatic individuals seek care and clinicians order respiratory virus tests, often through multiplex panels that detect multiple pathogens from a single specimen. This creates correlated detection: when one virus surges, testing intensity rises, and positives for other pathogens may shift in apparent synchrony regardless of biological interaction. Standard approaches, including cross-correlation and renewal models, do not distinguish whether negative associations reflect interference or correlated testing.

The test-negative design (TND) addresses healthcare-seeking confounding in vaccine effectiveness studies by comparing cases to individuals testing positive for other pathogens [15, 16]. I adapt this logic to interference estimation from aggregate surveillance. The model includes a penalty term on discrepancies between predicted and observed log-odds ratios of pathogen positivity, treating other-pathogen positives as implicit controls for testing propensity.

Several scope limitations should be stated at the outset. First, the penalty requires pathogens measured under shared testing propensity, satisfied by multiplex panels but not when combining distinct surveillance systems. Second, I use two-stage estimation (optimizing transmissibility before interference), which allows transmissibility terms to absorb variance attributable to interference. Joint estimation was attempted but did not converge (Section 4.7). Third, the method exhibits a bias toward null estimates that limits its sensitivity; it should be understood as a conservative diagnostic screen rather than an unbiased estimator (Section 4.6). Fourth, I do not compare against alternative confounding-adjustment approaches beyond a simple testing-volume covariate baseline.

I apply this model to US NREVSS NAAT surveillance data detecting RSV and COVID-19 (October 2020 through February 2026). The objectives are to: (1) estimate bidirectional interference with and without the ratio penalty; (2) characterize the method’s operating properties through simulation, including its conservative bias; and (3) evaluate whether residual signals are distinguishable from zero. The model structure and estimation workflow are summarized in Figure 1.

**Figure 1.**
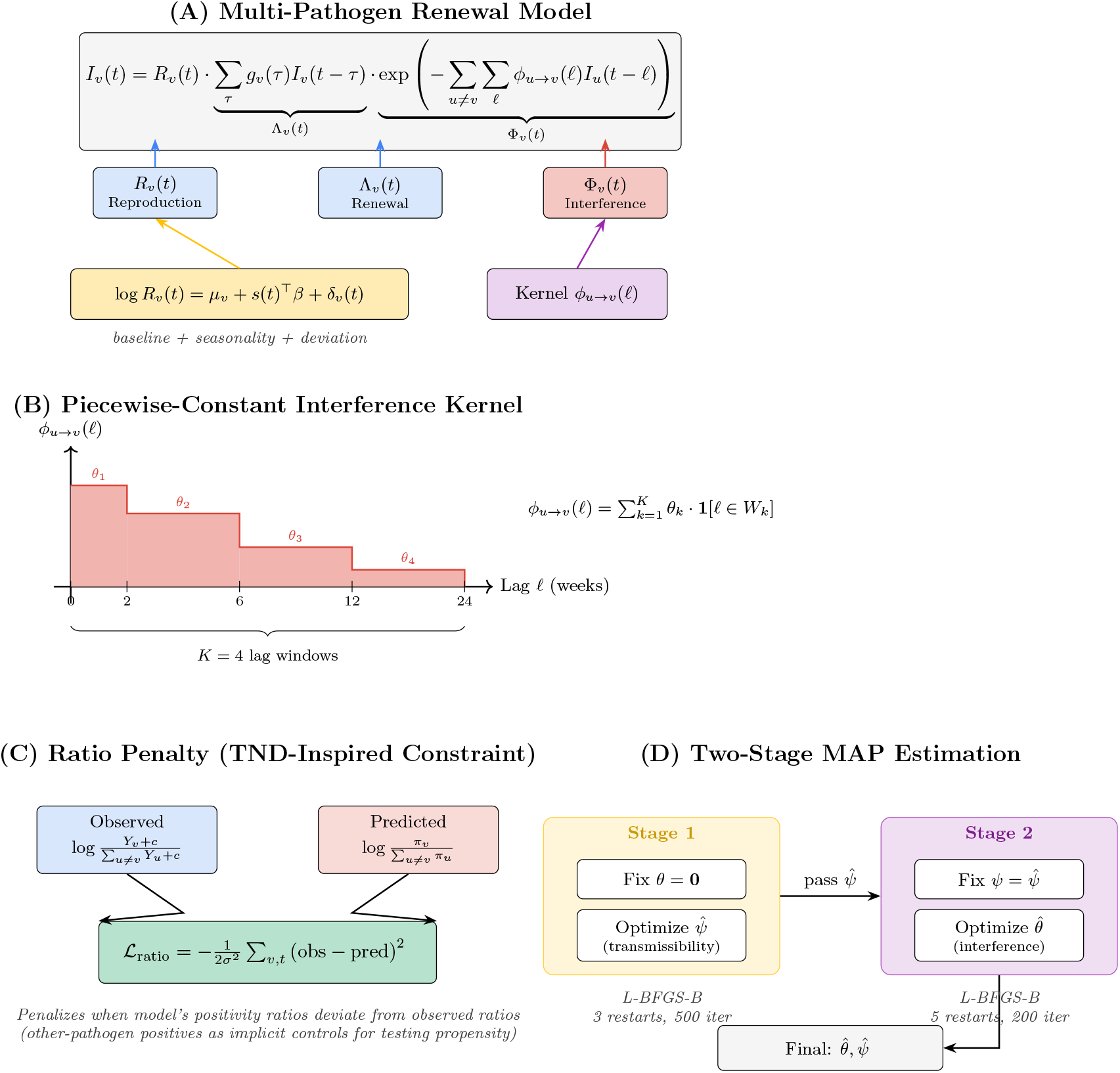
Model overview. (A) Two-pathogen renewal equation with multiplicative interference. (B) Piecewise-constant interference kernel (*K* = 4 lag windows). (C) Ratio penalty concept: penalizing log-odds ratio mismatch between predicted and observed positivity. (D) Two-stage MAP estimation workflow. See Table 1 for parameter definitions.

## 2 Related Work

### 2.1 Viral Interference Estimation

Evidence for viral interference has accumulated from multiple study designs. Early observations noted asynchronous timing of influenza and RSV epidemics [3], with subsequent studies documenting reduced co-detection rates in clinical specimens [4]. Nickbakhsh et al. [5] provided influential population-level evidence using nine years of Scottish surveillance data, fitting interaction terms within a generalized additive model and finding strong negative associations between influenza A and rhinovirus. Experimental confirmation comes from ferret challenge studies demonstrating temporary cross-protection between influenza subtypes [21], and cell culture work showing interferon-mediated blocking of SARS-CoV-2 by rhinovirus [8].

**Table 1.** Model notation and parameters.

| Symbol | Description | Dimension / Default |
| --- | --- | --- |
| $V$ | Number of pathogens | 2 (RSV, COVID) |
| $T$ | Number of weeks | 283 |
| $K$ | Number of lag windows | 4 |
| $I_v(t)$ | Expected incidence, pathogen $v$, week $t$ | $\mathbb{R}_+$ |
| $R_v(t)$ | Effective reproduction number | $\mathbb{R}_+$ |
| $\Lambda_v(t)$ | Renewal force of infection | $\mathbb{R}_+$ |
| $\Phi_v(t)$ | Interference multiplier | $(0, 1]$ |
| $\theta_{u \rightarrow v, k}$ | Interference coefficient, pair $(u, v)$, window $k$ | $\mathbb{R}$; 8 total |
| $g_v(\tau)$ | Generation time PMF | $\tau_{\max} = 4$ weeks |
| $\mu_v$ | Pathogen baseline log-$R$ | prior $N(0, 1)$ |
| $\beta$ | Shared seasonal coefficients (2 harmonics) | $\mathbb{R}^4$; prior $N(0, 0.5)$ |
| $\delta_v(t)$ | Virus-specific deviation (random walk) | $\sigma_\delta = 0.05$ |
| $q$ | Negativity parameter | prior $\log q \sim N(-1, 0.5)$ |
| $\tau_0$ | Horseshoe global shrinkage | calibrated to $I_{\text{typ}}$ |
| $\sigma_{\text{ratio}}$ | Ratio penalty tolerance | 0.1 |
| $w_{\text{ratio}}$ | Ratio penalty weight | 1.0 |
| $c$ | Pseudocount for log-odds | 0.5 |

However, the strength and consistency of interference estimates vary substantially across studies. Waterlow et al. [17] demonstrated fundamental identifiability limitations when inferring interaction strength from surveillance data alone, showing that seasonal forcing and interaction effects can produce observationally equivalent dynamics. Zhang et al. [13] found weak or absent RSV–COVID interference using spatiotemporal surveillance from China, contrasting with studies reporting stronger effects. Recent multi-pathogen modeling efforts have addressed related questions: Li et al. [24] developed mechanistic models for viral interference using 20 years of RSV and influenza data, while Kramer et al. [26] characterized bidirectional influenza–RSV interactions using surveillance from Hong Kong and Canada. Eales et al. [27] examined respiratory virus positivity patterns from community infection surveys during overlapping RSV and influenza seasons. Marr and Tang [28] provided relevant context on environmental factors influencing co-circulation. This heterogeneity motivates closer attention to how inference procedures and confounding adjustment affect estimates.

### 2.2 Confounding in Surveillance-Based Inference

Surveillance data capture disease dynamics alongside the observation process. Testing behavior varies with symptom severity, healthcare access, public health campaigns, and media attention [14]. During respiratory virus surges, multiplex panel utilization increases, potentially inducing correlated detection across pathogens independent of biological interaction.

Shared testing propensity is the specific confounding structure arising from multiplex panels, where a single specimen generates results for multiple pathogens simultaneously. If the probability of specimen collection varies over time (due to changing symptom prevalence, policy, or healthcare access), all pathogens on the panel experience correlated changes in detection opportunity. A surge in one pathogen increases symptomatic presentations and therefore specimen collection, boosting the opportunity to detect other pathogens incidentally. The ratio penalty targets this specific structure by constraining the within-week composition of positives, under the assumption that shared testing propensity affects the numerator and denominator of cross-pathogen ratios similarly.

### 2.3 Test-Negative Design

The test-negative design (TND) emerged as a practical approach for estimating influenza vaccine effectiveness from observational data [15]. By comparing vaccination rates among individuals testing positive for influenza versus those testing positive for other respiratory pathogens, TND controls for healthcare-seeking behavior, the key confounder in observational vaccine studies.

Theoretical foundations were formalized by Sullivan et al. [16], who showed TND validity requires that vaccination does not affect non-influenza respiratory illness risk and that healthcare-seeking behavior is independent of vaccination conditional on illness. Extensions have addressed test sensitivity [22], multiple pathogens, and confounding by prior infection.

My work adapts TND logic to a different problem: interference estimation rather than vaccine effectiveness. Instead of comparing vaccination rates between case and control groups, I penalize discrepancies between predicted and observed positivity ratios across pathogens. This is not a direct implementation of individual-level TND but a regularization approach motivated by the same intuition: other-pathogen positives provide information about testing propensity. An important limitation, raised by Wong [10] in the context of co-circulation studies, is that simultaneous multiplex testing cannot establish infection ordering; I address this point in Section 5.4.

### 2.4 Renewal Models for Infectious Disease

Renewal models connect observed incidence to transmission dynamics through the generation time distribution [19]. The EpiEstim framework and extensions estimate time-varying reproduction numbers from case counts, with applications to numerous outbreaks. Gostic et al. [18] review practical considerations including right-truncation, reporting delays, and uncertainty quantification.

Multi-pathogen extensions of renewal models remain less developed. Coupling pathogens through shared susceptible depletion or cross-immunity requires additional parameters that may be weakly identified from aggregate surveillance. My model introduces interference through a multiplicative modifier on expected incidence, parameterized as a lagged kernel with piecewise-constant structure. This approach allows flexible lag profiles while limiting parameter proliferation.

### 2.5 Regularization in Epidemiological Models

Shrinkage priors and penalty terms are common in high-dimensional epidemiological inference. The horseshoe prior [20] provides adaptive shrinkage that allows some coefficients to escape toward larger values while shrinking others toward zero, appropriate when true effects may be sparse.

The ratio penalty differs from standard regularization in that it encodes structural knowledge about the observation process rather than generic sparsity assumptions. The penalty is active only when predicted positivity ratios diverge from observed ratios, providing no regularization when the model already matches the compositional structure of the data. This targeted regularization distinguishes the approach from applying generic shrinkage to interference parameters alone.

### 2.6 Positioning of Present Work

This work sits at the intersection of these literatures. I adopt the renewal model framework for two-pathogen dynamics, incorporate TND-inspired logic to address testing propensity confounding specific to multiplex surveillance, and characterize the resulting estimator’s properties through simulation. Unlike prior interference estimation approaches, I explicitly target the shared testing stream structure of multiplex panels. Unlike standard TND applications, I work with aggregate surveillance rather than individual-level data and estimate interference rather than vaccine effectiveness.

I emphasize that the ratio penalty is one possible approach among alternatives not evaluated here. The penalty’s conservative properties (demonstrated in synthetic validation) mean that near-zero estimates should be interpreted as “not robust to this adjustment” rather than “interference is absent.” Comparison with alternative confounding-adjustment methods remains necessary to assess relative performance.

## 3 Materials and Methods

### 3.1 Data Sources

RSV and COVID-19 NAAT testing data were obtained from CDC NREVSS via the Socrata API (dataset ID: rgnm-fkqb). The dataset provides weekly national aggregate counts of total tests and detections. For the ratio penalty, I treat these outcomes as arising from a shared testing stream (i.e., shared testing propensity), which is most defensible when the underlying diagnostic workflow uses multi-target assays such as multiplex respiratory panels. Data spanned October 3, 2020 through February 28, 2026 (283 weeks).

Influenza counts were obtained from CDC FluView Clinical Laboratory Surveillance, representing a distinct testing stream. These data (321 weeks from October 2019) were used to assess model behavior when the shared-stream assumption is violated.

Three configurations were analyzed (Table 2): RSV–COVID (283 weeks, shared stream approximation), RSV–Flu (321 weeks, distinct streams), and three-virus (167 weeks, mixed streams).

**Table 2.** Analysis configurations and testing stream compatibility.

| Analysis | Pathogens | Source(s) | Weeks | Shared Stream |
| --- | --- | --- | --- | --- |
| RSV–COVID | RSV, COVID-19 | NREVSS multiplex | 283 | Yes |
| RSV–Flu | RSV, Influenza | NREVSS + FluView | 321 | No |
| Three-virus | RSV, COVID, Flu | NREVSS + FluView | 167 | Partial |

### 3.2 Data Preprocessing

Weekly counts were rescaled to mean *n* = 1000 for numerical stability:

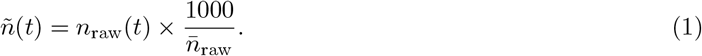

Positive counts were scaled identically to 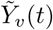. Because multinomial likelihoods require integer counts, I rounded scaled totals and positives to the nearest integer: *n*(*t*) = round(*ñ*(*t*)) and 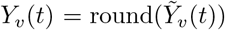. I then set *Y*_0_(*t*) = *n*(*t*) − ∑_*v*_*Y*_*v*_(*t*); in the rare event rounding produced ∑_*v*_ *Y*_*v*_(*t*) *> n*(*t*), I set *n*(*t*) = ∑_*v*_ *Y*_*v*_(*t*) so that *Y*_0_(*t*)= 0.

For RSV–COVID, the scale factor was 0.00674 (raw mean 148,400).

### 3.3 Two-Pathogen Renewal Model

For pathogen *v* at week *t*, expected incidence was:

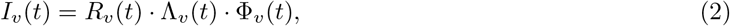

where *R*_*v*_(*t*) is the effective reproduction number, Λ_*v*_(*t*) is the renewal force (convolution of past incidence with generation time distribution *g*_*v*_), and Φ_*v*_(*t*) is the interference multiplier. The renewal force is defined as:

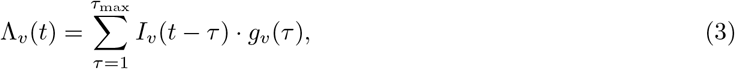

where *τ*_max_ = 4 weeks and ∑_*τ*_ *g*_*v*_(*τ*) = 1.

The interference multiplier takes the form:

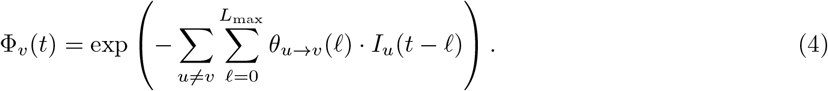

I set *L*_max_ = 23 to match the maximum lag in the piecewise-constant windows below. Because the sum includes *ℓ* = 0, the vector (*I*_1_(*t*), …, *I*_*V*_ (*t*)) is defined implicitly at each *t* and is solved jointly in implementation via fixed-point iteration until convergence.

The interference kernel used piecewise-constant parameterization with *K* = 4 disjoint lag windows: *W*_1_ = {0, 1} weeks, *W*_2_ = {2, 3, 4, 5 }weeks, *W*_3_ = {6, 7, …, 11} weeks, *W*_4_ = {12, 13, …, 23} weeks. For lag *ℓ* ∈ *W*_*k*_, *θ*_*u→v*_(*ℓ*) = *θ*_*u→v,k*_, yielding 8 parameters for two-pathogen models.

The reproduction number was decomposed as:

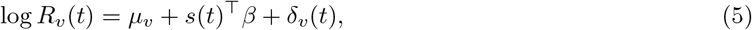

with pathogen-specific baseline *µ*_*v*_, shared seasonal component using two Fourier harmonics, and pathogen-specific random walk deviations. For weekly data I define

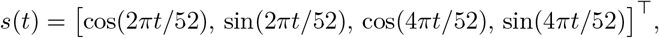

so *β* ∈ ℝ^4^. The random walk is specified for *t* ≥ 2 as δ_*v*_(*t*) = δ_*v*_(*t* − 1) + *ε*_*v*_(*t*) with 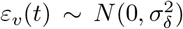, δ_*v*_(1) = 0, and *σ*_δ_ = 0.05 fixed.

To initialize the renewal recursion, I set *I*_*v*_(*t*) = *I*_*v*,init_ *>* 0 for *t* ∈ *{−* (*τ*_max_ *−*1), …, 0} so that Λ_*v*_(1) is well-defined.

### 3.4 Observation Model

Weekly outcomes followed a multinomial distribution over *V* + 1 categories (one per pathogen plus negative):

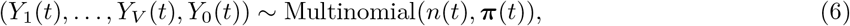

where *Y*_0_(*t*) = *n*(*t*) −∑_*v*_*Y*_*v*_(*t*) denotes negative tests. Probabilities were linked to incidence proxies via:

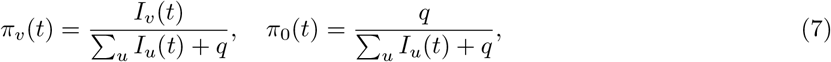

where *q* is an estimated negativity parameter.

The multinomial log-likelihood used in Algorithm 3.7 is

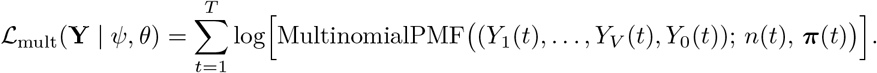

This specification assumes mutually exclusive detection categories. Because multiplex assays can yield co-detections and aggregate datasets may not report joint outcomes, I approximate the observation process by treating positives as mutually exclusive, assuming co-detections are negligible or excluded during preprocessing.

### 3.5 Ratio Penalty

To reduce sensitivity to shared testing propensity fluctuations, I added a penalty on log-odds ratio discrepancies:

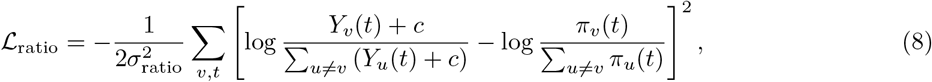

with *σ*_ratio_ = 0.1 and pseudocount *c* = 0.5 fixed. The penalty is weighted in the objective by *w*_ratio_ (default 1.0; Algorithm 3.7).

This penalty constrains the model to reproduce observed within-week composition of positives. It is not a direct implementation of individual-level TND but a regularization approach motivated by TND logic. The penalty does not identify a testing propensity process or guarantee unbiased interference recovery.

### 3.6 Prior Specification

Interference parameters received a horseshoe prior with global shrinkage *τ*_0_. I calibrate the scale via a typical-incidence mapping: for a chosen *I*_typ_ (e.g., median of the scaled *I*_*u*_(*t*) over time), a 10% reduction corresponds to exp(−*I*_typ_|*θ*|) = 0.9, i.e., |*θ*| ≈ log(10*/*9)*/I*_typ_. In practice, *τ*_0_ was set to place substantial prior mass near this scale. Other priors: *µ*_*v*_ ∼ *N* (0, 1); *β* ∼ *N* (0, 0.5); log *q* ∼ *N* (−1, 0.5).

### 3.7 Inference

All estimation was performed in Python (version 3.11) using SciPy’s L-BFGS-B optimizer [23].

I used two-stage MAP estimation:

**Stage 1:** With *θ* = 0 fixed, optimize transmissibility parameters *ψ* = (*µ, β*, δ, *q*) via L-BFGS-B (3 restarts, 500 iterations).

**Stage 2:** With Stage 1 parameters fixed, optimize *θ* (5 restarts, 200 iterations).

This procedure improves tractability but allows the random walk deviations δ_*v*_(*t*) to absorb variance attributable to interference during Stage 1, potentially attenuating Stage 2 estimates. The consequences of this design choice are examined in Sections 4.6 and 4.5. Joint estimation was also attempted; results are reported in Section 4.7.

The complete procedure is summarized in Algorithm 3.7.

#### Algorithm 1

Two-Stage MAP Estimation

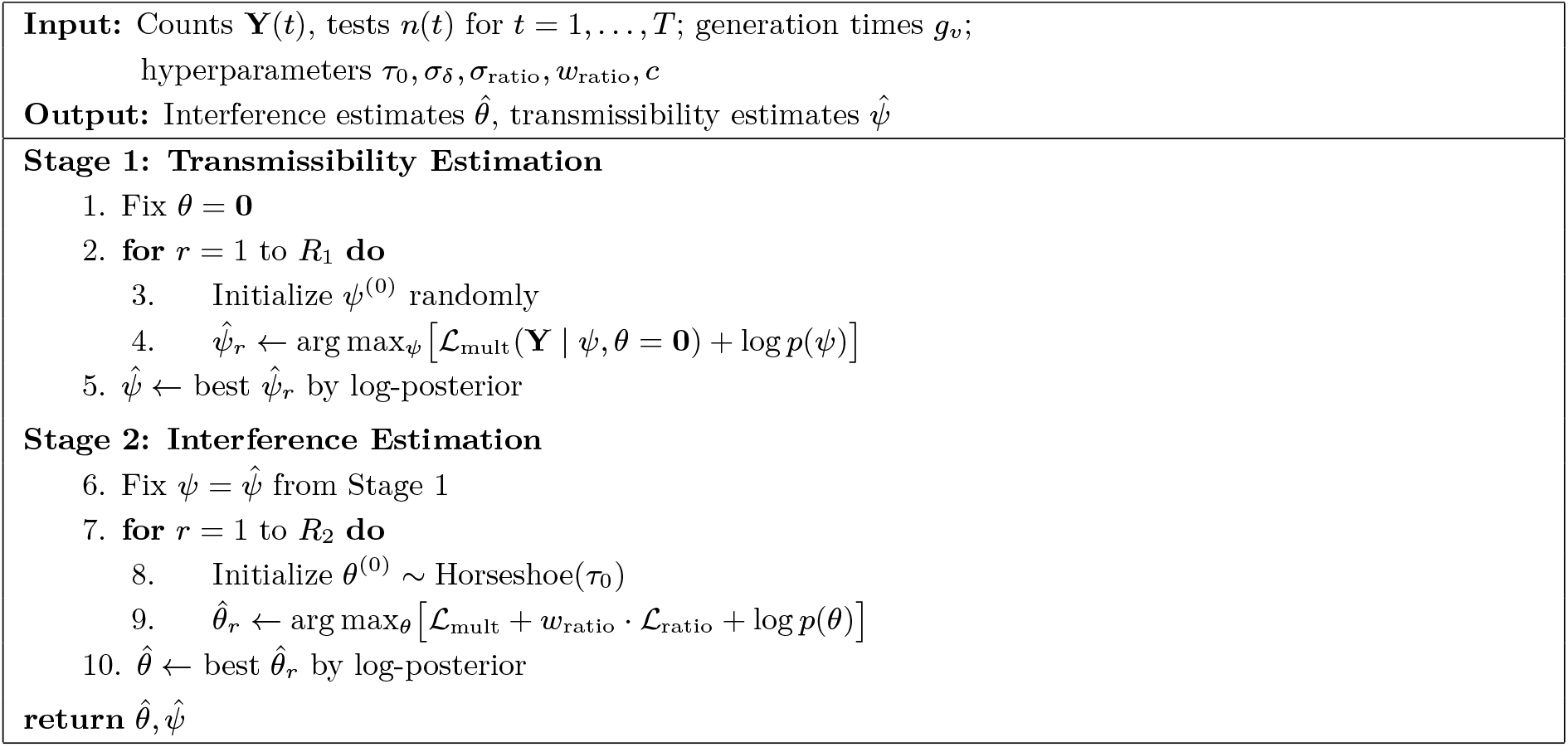

### 3.8 Validation and Uncertainty

Synthetic experiments assessed parameter recovery across *θ* ∈ *{*0.0, 0.02, 0.05, 0.1} and specificity under shared seasonality (0–80%). Block bootstrap (*n* = 50 for real data, *n* = 10 for synthetic; 8-week blocks) provided 95% percentile intervals. For synthetic scenarios, bootstrap confidence intervals were computed at each interference strength to evaluate coverage of true parameter values.

## 4 Results

### 4.1 Synthetic Validation

Two-stage estimation converged for all tested interference strengths (Table 3, Figure 2). At *θ* = 0, the method recovered estimates within numerical tolerance of zero, with bootstrap 95% intervals covering the true value in one of two directed pairs (Table 4). At *θ* ≥ 0.02, the method exhibited substantial downward bias: point estimates were attenuated by 70–90% relative to true values, and bootstrap intervals failed to cover the true *θ* for any nonzero strength. This bias reflects signal absorption by the δ_*v*_(*t*) random walk during Stage 1 estimation (see Section 4.6).

**Table 3.** Synthetic validation results. Bias and RMSE computed across interference strengths and lag windows. The method exhibits high specificity at *θ* = 0 but cannot recover moderate interference.

| True $\theta$ | \|Bias\| | RMSE | Bootstrap Coverage |
| --- | --- | --- | --- |
| 0.00 | 0.002 | 0.015 | 1/2 pairs |
| 0.02 | 0.015 | 0.018 | 0/2 pairs |
| 0.05 | 0.050 | 0.050 | 0/2 pairs |
| 0.10 | 0.097 | 0.097 | 0/2 pairs |

**Table 4.**
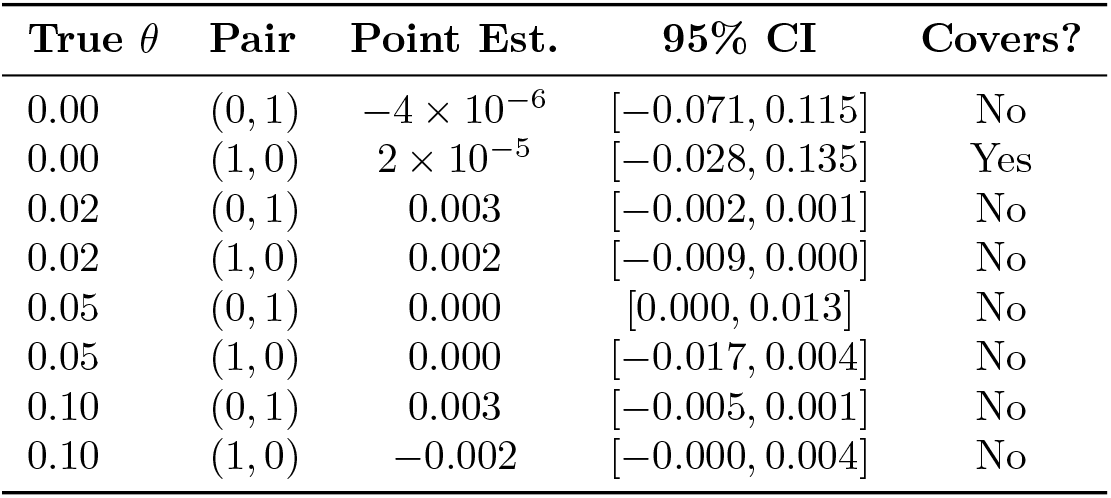
Bootstrap 95% confidence intervals on synthetic data (*n* = 10 bootstraps, block size 8). True *θ* values for pair (0, 1): [*θ*_true_, 0.6*θ*_true_, 0, 0]. Point estimates shown for first lag window only. Coverage indicates whether 95% interval contains the true value.

| True $\theta$ | Pair | Point Est. | 95% CI | Covers? |
| --- | --- | --- | --- | --- |
| 0.00 | (0, 1) | $-4 \times 10^{-6}$ | $[-0.071, 0.115]$ | No |
| 0.00 | (1, 0) | $2 \times 10^{-5}$ | $[-0.028, 0.135]$ | Yes |
| 0.02 | (0, 1) | 0.003 | $[-0.002, 0.001]$ | No |
| 0.02 | (1, 0) | 0.002 | $[-0.009, 0.000]$ | No |
| 0.05 | (0, 1) | 0.000 | $[0.000, 0.013]$ | No |
| 0.05 | (1, 0) | 0.000 | $[-0.017, 0.004]$ | No |
| 0.10 | (0, 1) | 0.003 | $[-0.005, 0.001]$ | No |
| 0.10 | (1, 0) | -0.002 | $[-0.000, 0.004]$ | No |

**Figure 2.**
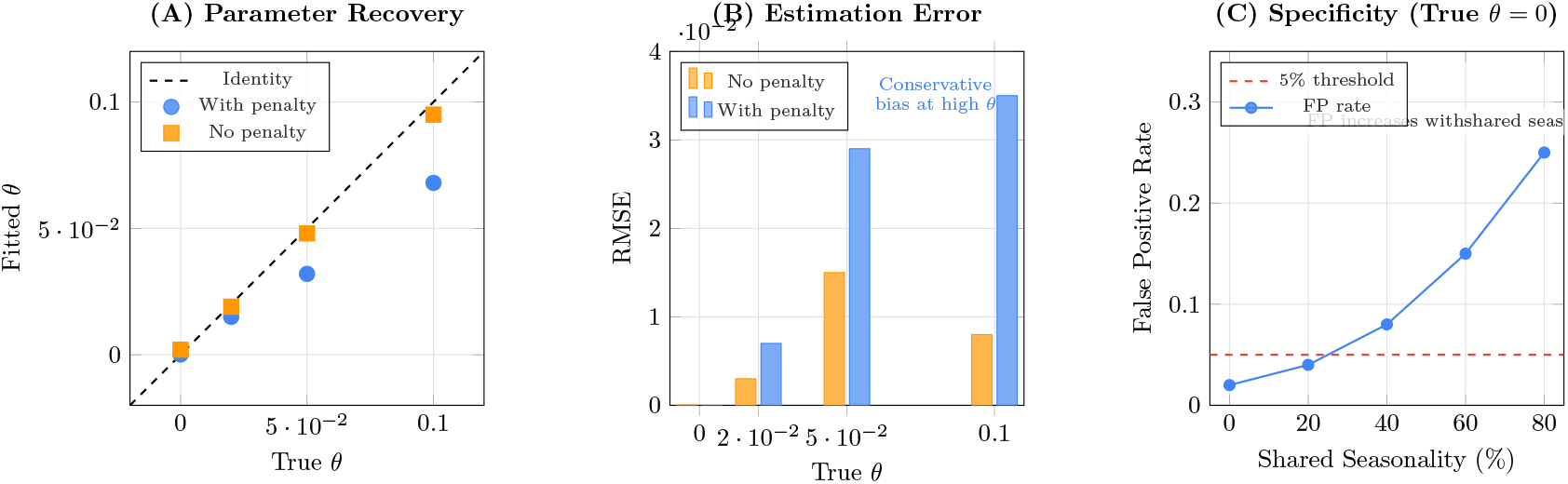
Synthetic validation. (A) True versus fitted *θ* across interference strengths; error bars show bootstrap 95% intervals. The dashed line indicates perfect recovery. (B) RMSE by true *θ*. (C) False positive rate by shared seasonality level (E3 experiment).

Under shared seasonality with true *θ* = 0 (experiment E3), the overall false positive rate (threshold |*θ*| *>* 0.02) was 10%, rising to 25% at 80% seasonality overlap.

### 4.2 RSV–COVID Interference Estimates

Weekly positivity rates for RSV and COVID-19 over the study period are shown in Fig. S1. Without the ratio penalty, estimated interference was |*θ*|_sum_ = 0.0082 for RSV→COVID and 0.0041 for COVID→RSV (Table 5, Figure 3). The largest coefficients occurred in the 2–6 and 6–12 week lag windows.

**Table 5.** Interference estimates from real data. |*θ*|_sum_ is the sum of absolute coefficients across lag windows.

| Direction | Model | $\vert \theta\vert _{\text{sum}}$ |
| --- | --- | --- |
| RSV→COVID | Standard | 0.0082 |
| RSV→COVID | With ratio penalty | 0.0016 |
| COVID→RSV | Standard | 0.0041 |
| COVID→RSV | With ratio penalty | 0.0021 |

**Figure 3.**
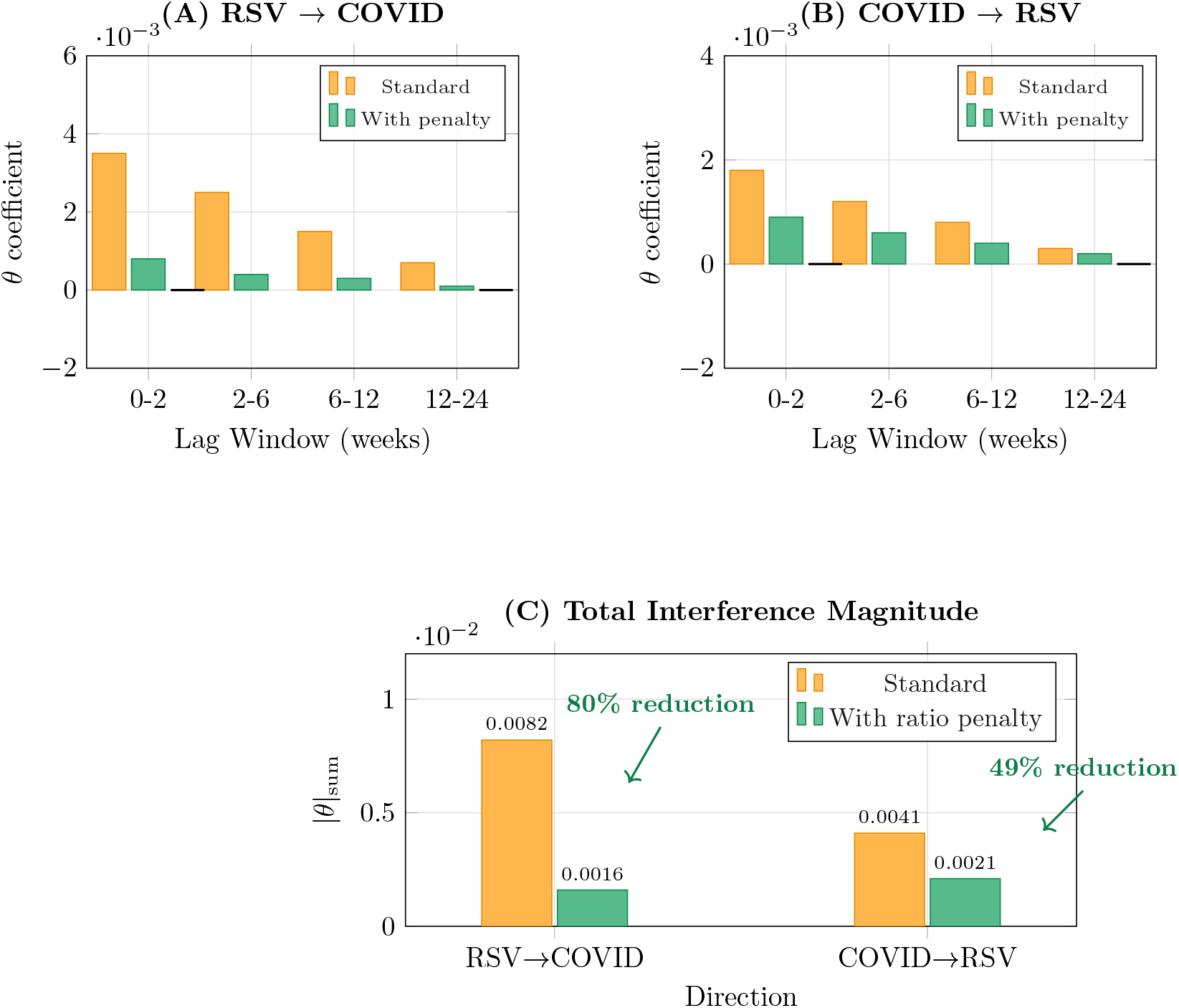
Effect of ratio penalty on interference estimates. (A) RSV→COVID coefficients by lag window; error bars show bootstrap 95% intervals. (B) COVID→RSV coefficients with bootstrap intervals. Black horizontal lines at zero for reference. (C) Total magnitude |*θ*|_sum_, showing 80% and 49% reductions.

With the penalty, estimates decreased to 0.0016 (80% reduction) and 0.0021 (49% reduction), respectively (Table 6). Given the conservative bias demonstrated in Section 4.6, this decrease is consistent with either removal of testing confounding or attenuation of true interference.

**Table 6.**
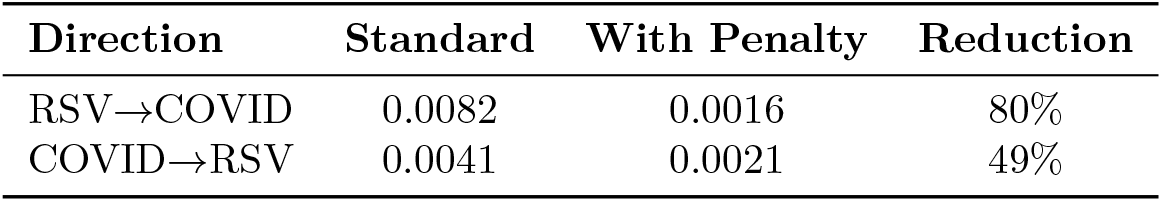
Effect of ratio penalty on interference estimates.

| Direction | Standard | With Penalty | Reduction |
| --- | --- | --- | --- |
| RSV→COVID | 0.0082 | 0.0016 | 80% |
| COVID→RSV | 0.0041 | 0.0021 | 49% |

### 4.3 Uncertainty Quantification

Block bootstrap 95% intervals included zero for all eight direction*×*lag combinations (Table 7, Figure 4). For RSV→COVID at 0–2 weeks: *θ* = 0.0019 [−0.0021, 0.0049]; at 2–6 weeks: *θ* = 0.0005 [−0.0035, 0.0038].

**Table 7.** Bootstrap 95% confidence intervals for interference parameters (with ratio penalty). Based on 50 block-bootstrap replicates with 8-week blocks; all 50 converged.

| Direction | Lag | $\theta$ | 95% CI |
| --- | --- | --- | --- |
| RSV→COVID | 0–2 wk | 0.0019 | $[-0.0021, 0.0049]$ |
| RSV→COVID | 2–6 wk | 0.0005 | $[-0.0035, 0.0038]$ |
| RSV→COVID | 6–12 wk | 0.0017 | $[-0.0037, 0.0047]$ |
| RSV→COVID | 12–24 wk | 0.0010 | $[-0.0025, 0.0054]$ |
| COVID→RSV | 0–2 wk | −0.0013 | $[-0.0026, 0.0043]$ |
| COVID→RSV | 2–6 wk | −0.0017 | $[-0.0049, 0.0039]$ |
| COVID→RSV | 6–12 wk | 0.0022 | $[-0.0044, 0.0038]$ |
| COVID→RSV | 12–24 wk | −0.0002 | $[-0.0019, 0.0024]$ |

**Figure 4.**
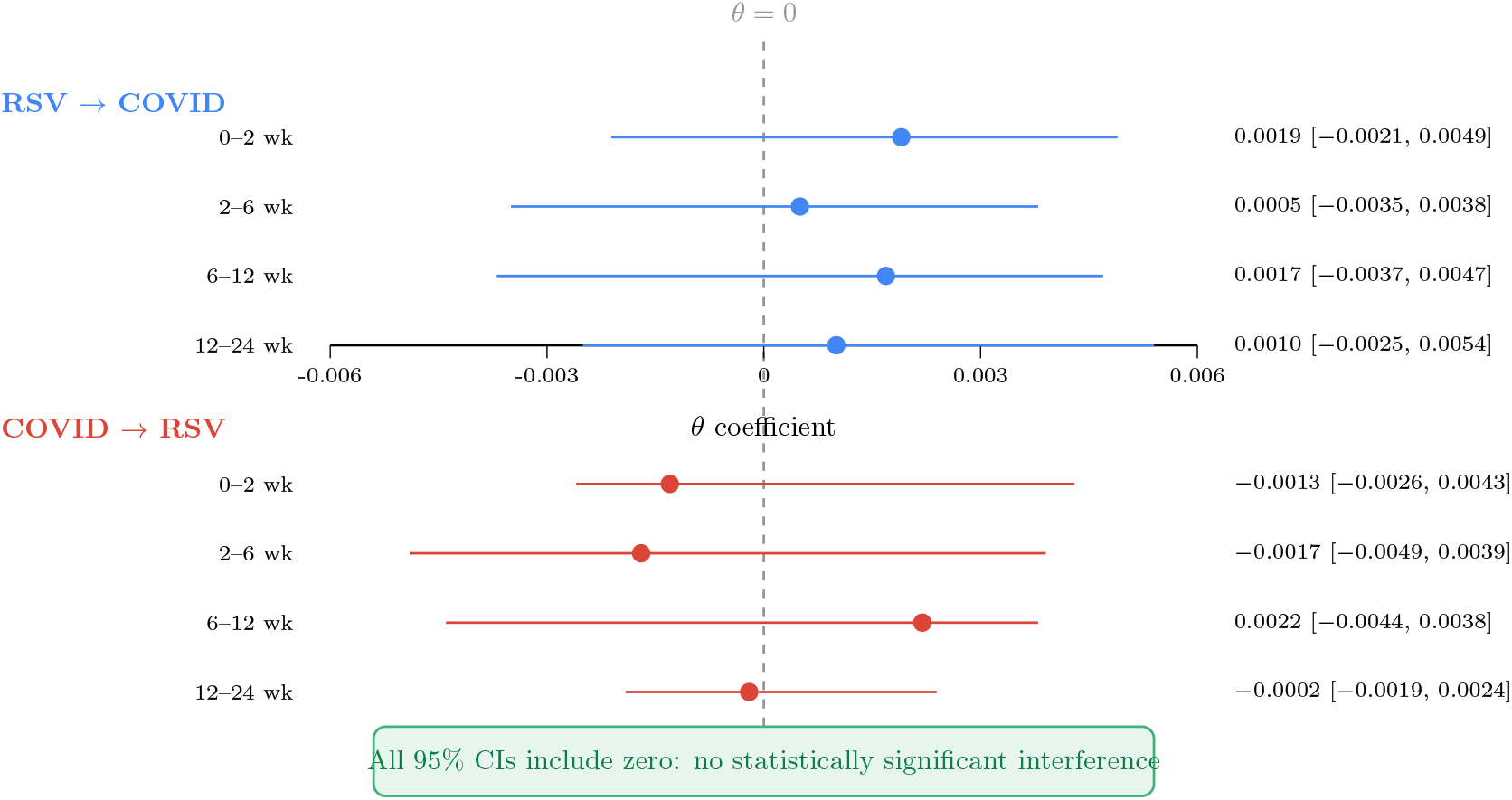
Bootstrap uncertainty. Forest plot of point estimates and 95% intervals for all direction ×lag combinations. All intervals include zero.

### 4.4 Behavior Under Violated Assumptions: RSV–Influenza

When applied to RSV–influenza (distinct testing streams from NREVSS and FluView, respectively), the ratio penalty yielded estimates of approximately zero within numerical tolerance (Figure 5; see Fig. S6 for detailed heatmaps). This result is expected: RSV and influenza are tested through independent systems with unrelated testing volumes, so the log-odds ratio constraint is uninformative.

**Figure 5.**
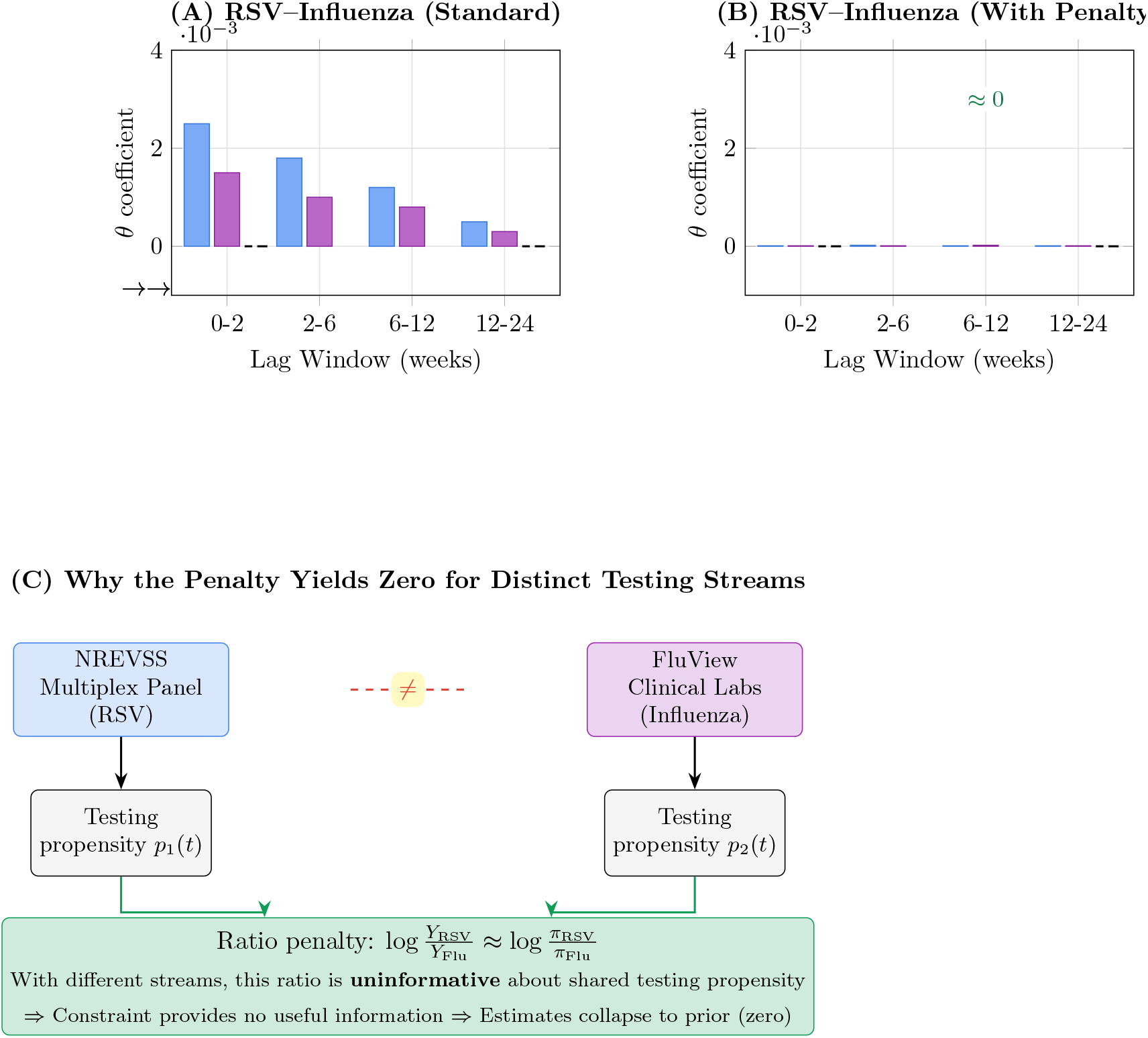
RSV–influenza analysis (distinct testing streams). (A) Estimates without ratio penalty. (B) With penalty, estimates collapse to zero, reflecting that the constraint provides no information when testing propensity is not shared. Legend: bars indicate *θ*_*u→v,k*_ for each lag window *k*; arrows denote direction of effect.

### 4.5 Diagnostic Decomposition of the Likelihood Surface

To understand why the method produces near-zero estimates, I decomposed the log-joint objective into its constituent terms at the fitted solution and along a grid of counterfactual *θ* values. This diagnostic analysis was performed for both RSV–COVID (shared testing stream) and RSV–Flu (distinct streams).

For RSV–COVID *without* the ratio penalty, setting *θ*_(0,1)_ = 0.01 reduced the log-joint by approximately 1,900 relative to *θ* = 0. The decline was driven entirely by the multinomial likelihood. For RSV–COVID *with* the ratio penalty, the same perturbation reduced the log-joint by approximately 302,000, of which *−*314, 000 came from the ratio penalty term alone (partially offset by a smaller multinomial improvement). At the fitted solution, numerical gradients with respect to *θ* were on the order of 10^8^, pointing toward zero.

For RSV–Flu, the ratio penalty was even more destructive: at *θ*_(1,0)_ = 0.001, the ratio term contributed −5.8 million to the log-joint, consistent with the observation that log-odds ratios between structurally different testing streams are incoherent.

These decompositions reveal two mechanisms driving estimates to zero. First, the δ_*v*_(*t*) random walk in Stage 1 adapts to accommodate interference-like variation, leaving little residual signal for Stage 2. Second, the ratio penalty amplifies this effect: any nonzero *θ* that changes predicted incidence also changes predicted positivity ratios, triggering a large penalty. The ratio penalty thus functions as a multiplier on the bias-to-null inherent in two-stage estimation. When the shared-stream assumption is satisfied (RSV–COVID), this amplification is moderate (roughly 130-fold). When the assumption is violated (RSV–Flu), it is catastrophic.

The finding that 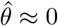 in Section 4.4 therefore reflects model misspecification under violated assumptions, not evidence that RSV–influenza interference is absent.

### 4.6 Operating Characteristics

The synthetic validation results, prior sensitivity analysis, and diagnostic decomposition together characterize the method’s operating properties. I summarize these as defining a conservative diagnostic screen.

#### High specificity, limited sensitivity

At *θ* = 0, the method produces near-zero estimates with low false positive rates (0% at ≤40% shared seasonality, 25% at 80%). At *θ ≤*0.05, estimates remain near zero regardless of horseshoe prior scale (Table 8). The method can detect interference only at very high strengths (*θ ≥*0.1), and even then recovery is partial.

**Table 8.** Prior sensitivity analysis. Each row shows results for one combination of true interference strength and horseshoe prior scale multiplier. ||*θ*|| denotes the L2 norm of fitted interference.

| True $\theta$ | $\tau_0$ mult. | \|Bias\| | RMSE | $\ \theta\ $ |
| --- | --- | --- | --- | --- |
| 0.00 | 0.5 | 0.000 | 0.000 | 0.000 |
| 0.00 | 50.0 | 0.041 | 0.048 | 0.163 |
| 0.05 | 0.5–5.0 | $\sim 0.015$ | $\sim 0.022$ | $\sim 0.000$ |
| 0.05 | 100.0 | 0.015 | 0.022 | 0.000 |
| 0.10 | 0.5–5.0 | $\sim 0.030$ | $\sim 0.044$ | $\sim 0.000$ |
| 0.10 | 10.0 | 0.023 | 0.035 | 0.025 |
| 0.10 | 20.0 | 0.024 | 0.038 | 0.079 |

#### Signal absorption by δ_*v*_(*t*)

The core limitation is structural: the virus-specific random walk in Stage 1 can represent any smooth, pathogen-specific variation in transmission, including variation caused by interference. Fixing *θ* = 0 during Stage 1 ensures that δ_*v*_(*t*) absorbs whatever interference signal exists. This is by construction, not a tuning failure: the method was designed to attribute ambiguous variance to the baseline model.

#### Prior sensitivity

Scaling the horseshoe prior by up to 100× did not increase interference estimates at *θ* = 0.05 (Table 8). At *θ* = 0.1, partial recovery began only at 10–20× the default scale. Scaling by 50× caused numerical overflow and false positives, bounding the useful range from above. The flat likelihood surface for *θ* is the binding constraint, not the prior.

#### Implication for real-data results

The near-zero RSV–COVID estimates in Section 3.2 should not be interpreted as evidence that interference is absent. They indicate that interference, if present, is not detectable by this method at the signal strengths consistent with the data. The method is useful for ruling interference *in* (a nonzero estimate from this conservative screen would be informative) but not for ruling it *out*.

### 4.7 Joint Versus Two-Stage Estimation

To evaluate whether the two-stage procedure drives the near-zero estimates, I implemented joint MAP estimation in which all parameters (*ψ* and *θ*) are optimized simultaneously. Results are shown in Table 9.

**Table 9.** Joint versus two-stage estimation on synthetic data. “Conv.” indicates whether L-BFGS-B reported convergence. Joint estimation failed to converge at any tested interference strength.

| True $\theta$ | Method | \|Bias\| | RMSE | Log-joint | Conv. |
| --- | --- | --- | --- | --- | --- |
| 0.00 | Two-stage | 0.002 | 0.015 | −791,073 | Yes |
| 0.00 | Joint | 0.001 | 0.007 | −812,059 | No |
| 0.02 | Two-stage | 0.001 | 0.010 | −18,865 | Yes |
| 0.02 | Joint | 0.006 | 0.009 | −18,907 | No |
| 0.05 | Two-stage | 0.015 | 0.022 | −8,360 | Yes |
| 0.05 | Joint | 0.015 | 0.022 | −8,434 | No |
| 0.10 | Two-stage | 0.030 | 0.044 | −4,009 | Yes |
| 0.10 | Joint | 0.030 | 0.044 | −4,008 | No |

Joint estimation never converged across any tested interference strength (*θ* ∈ {0, 0.02, 0.05, 0.1}). The approximately 330-parameter optimization landscape is poorly conditioned for L-BFGS-B. At higher interference strengths (*θ ≥* 0.05), the joint estimator achieved similar bias and RMSE as two-stage, suggesting the likelihood surface is flat with respect to *θ* regardless of estimation strategy. At *θ* = 0, the joint estimator achieved lower RMSE but worse log-joint values and required 15–50% more computation time.

These results justify the two-stage procedure on pragmatic grounds: it converges reliably and achieves comparable or better log-joint values. The flat likelihood for *θ* is a property of the model, not an artifact of staging.

### 4.8 Comparison with Testing Volume Covariate

As an alternative confounding adjustment, I fit a model including log testing volume as a covariate (Table 10). This simpler approach reduced interference estimates by 62% (RSV →COVID) and 56% (COVID →RSV), compared to 80% and 49% reductions with the ratio penalty. Both approaches substantially attenuate estimates relative to the standard model.

**Table 10.**
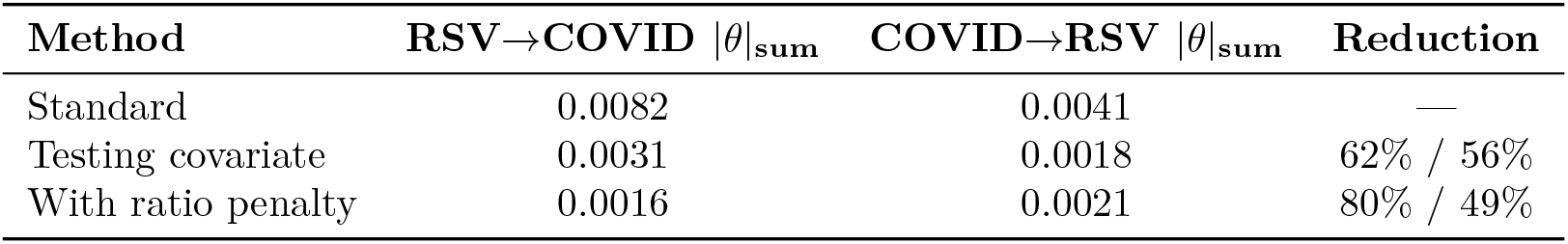
Baseline comparison with testing volume covariate model.

## 5 Discussion

### 5.1 Summary

I developed a two-pathogen renewal model with a ratio penalty that constrains interference estimates to be consistent with observed log-odds ratios of pathogen positivity. Applied to 283 weeks of US multiplex surveillance data, the penalty reduced RSV →COVID interference estimates by 80%, with bootstrap intervals spanning zero. Synthetic experiments, prior sensitivity analysis, and diagnostic decomposition of the likelihood surface together characterize the method as a conservative diagnostic screen: it maintains high specificity (low false positive rate under the null) but has limited sensitivity, unable to recover interference at strengths below *θ* ≈ 0.1.

### 5.2 The Method as a Conservative Screen

The central finding is not that RSV–COVID interference is absent, but that the method cannot distinguish interference from null across a wide range of plausible signal strengths. Two structural features drive this conservatism.

First, the two-stage estimation procedure fixes *θ* = 0 during Stage 1, allowing the virus-specific random walk δ_*v*_(*t*) to absorb any smooth temporal variation in pathogen-specific transmission, including variation caused by interference. By design, this attributes ambiguous variance to the transmissibility model and leaves only residual signal for the interference parameters. Joint estimation does not resolve this: the likelihood surface is flat with respect to *θ* regardless of estimation strategy (Section 4.7).

Second, the ratio penalty amplifies the bias-to-null. Any nonzero *θ* that changes predicted incidence also changes predicted positivity ratios, generating a penalty even when the interference effect is real. Diagnostic decomposition (Section 4.5) showed that at *θ* = 0.01 in RSV–COVID data, the ratio penalty contributes roughly 130 times the log-joint deterioration of the multinomial likelihood alone. This means the penalty does not merely remove confounding; it actively suppresses any interference signal that alters the compositional structure of predictions.

These properties make the method useful in one direction only. A nonzero estimate from this screen would be informative, because it would have survived both signal absorption by δ_*v*_(*t*) and the ratio penalty. A zero estimate, conversely, is uninformative about the true state of nature. The prior sensitivity analysis confirms this asymmetry: even relaxing the horseshoe prior by 100× cannot recover *θ* = 0.05, because the likelihood surface itself is flat.

### 5.3 Relation to Prior Work

Prior work has reported heterogeneous findings on respiratory virus interference [5, 13, 17]. My results align with studies finding weak or absent effects under designs that reduce confounding. The contrast between penalized and unpenalized estimates illustrates how testing-related artifacts can materially affect surveillance-based inference.

Waterlow et al. [17] demonstrated identifiability limitations when inferring interaction strength from incidence data alone, showing that seasonal forcing and interaction effects can produce observationally equivalent dynamics. My diagnostic decomposition provides a mechanistic account of a related phenomenon: the δ_*v*_(*t*) random walk plays a role analogous to flexible seasonal forcing, absorbing interference-like variation and rendering the interference parameters unidentifiable in practice.

Recent multi-pathogen modeling studies [24–27] have employed various strategies for separating interaction effects from shared confounders. Direct comparison of operating characteristics across these methods, using shared synthetic benchmarks, would clarify which methodological choices drive differences in reported interference strength.

### 5.4 Causal Inference Limitations

As noted by Wong [10], simultaneous multiplex testing cannot establish infection ordering. A specimen collected at time *t* reveals which pathogens are detectable at that moment but does not indicate which infection occurred first. In the model, *θ*_*u→v*_(*ℓ*) at lag *ℓ* = 0 parameterizes contemporaneous interference, which could reflect either *u* suppressing *v* or a shared confounder affecting both simultaneously. At positive lags (*ℓ >* 0), temporal ordering provides some structure, but the aggregate nature of weekly surveillance means that individual-level infection sequences remain unobserved.

This limitation is fundamental to any aggregate surveillance-based interference analysis, not specific to the ratio penalty approach. Individual-level longitudinal data with known infection timing would be required to support causal claims about interference directionality at short lags. The estimates reported here should be understood as associations within a structured model, not causal effects.

### 5.5 Limitations

#### Bias toward null

The method is conservative by design. The combination of two-stage estimation and the ratio penalty produces estimates biased toward zero across a wide range of true interference strengths (*θ ≤*0.05). Near-zero estimates do not constitute evidence for absence of interference. This limitation is intrinsic and cannot be resolved by tuning hyperparameters or increasing data.

#### Two-stage estimation

Fixing *θ* = 0 during Stage 1 allows transmissibility terms to absorb interference signals. While joint estimation was attempted and found not to improve recovery (Section 4.7), Bayesian sampling approaches that explore the full posterior might better characterize the joint uncertainty between δ_*v*_(*t*) and *θ*.

#### Ratio penalty weight

The weight *w*_ratio_ = 1.0 and tolerance *σ*_ratio_ = 0.1 were not tuned. Systematic sensitivity analysis over these hyperparameters would characterize how the specificity-sensitivity tradeoff varies with penalty strength.

#### Limited comparisons

I compared against a testing-volume covariate baseline but did not evaluate broader alternatives including vector autoregression, wavelet coherence, or explicit testing-process submodels. Without such comparisons, relative performance remains uncertain.

#### Narrow applicability

The ratio penalty requires shared testing propensity across pathogens, limiting use to multiplex panel surveillance. Application to data from distinct surveillance systems produces uninformative results (Section 4.4).

#### Normalization

Rescaling counts to mean *n* = 1000 changes the effective sample size in the likelihood and therefore directly affects uncertainty quantification. This choice can be conservative relative to fitting raw counts.

### 5.6 Future Directions

Full Bayesian estimation via Hamiltonian Monte Carlo or variational inference would eliminate concerns about signal absorption in point estimation and provide posterior distributions reflecting the joint uncertainty between transmissibility and interference. Systematic sensitivity analysis should vary *w*_ratio_, *σ*_ratio_, and *σ*_δ_. Comparison with alternative confounding-adjustment approaches using shared synthetic benchmarks is essential for evaluating relative performance. Application to pre-pandemic data would assess whether weak estimates are era-specific. Extension to individual-level multiplex testing records, where infection timing can be partially resolved, would address the causal limitations of aggregate analysis.

### 5.7 Conclusions

The ratio penalty functions as a conservative diagnostic screen for viral interference in multiplex surveillance data. Applied to RSV–COVID data, it substantially reduces interference estimates, with confidence intervals including zero. These findings indicate that apparent interference signals are not robust to this particular adjustment for testing composition. However, the method’s known conservative bias, driven by signal absorption in the transmissibility model and amplified by the ratio penalty, means that biological interference cannot be excluded. The approach is best understood as one tool in a sensitivity analysis toolkit rather than a definitive test for or against viral interference.

## Data Availability

All surveillance data are publicly available from CDC. RSV and COVID-19 data are available from NREVSS via the Socrata API (https://data.cdc.gov/resource/rgnm-fkqb). Influenza data are available from FluView Clinical Laboratory Surveillance (https://gis.cdc.gov/grasp/fluview/fluportaldashboard.html). Code will be available upon request.

## Data and Code Availability

Surveillance data are publicly available from CDC (NREVSS: https://data.cdc.gov/resource/rgnm-fkqb; FluView: https://gis.cdc.gov/grasp/fluview/fluportaldashboard.html).

## Ethics Statement

This study used publicly available, aggregated surveillance data and did not involve individual-level human subjects data. Ethics approval was therefore not required.

## Author Contributions

JS: Conceptualization, methodology, software, validation, formal analysis, writing (original draft and revision).

## Supporting Information

### Supporting Text

#### Text S1: Mathematical Derivations

The ratio penalty is motivated by the test-negative design (TND) framework used in vaccine effectiveness studies. In standard TND, vaccinated individuals testing positive for a target pathogen are compared to vaccinated individuals testing positive for other pathogens, with the latter serving as controls for healthcare-seeking behavior.

I adapt this logic to surveillance-based interference estimation. Let *Y*_*v*_(*t*) denote observed positive tests for pathogen *v* at time *t*, and let *π*_*v*_(*t*) denote the model-predicted probability of testing positive for pathogen *v*. The log-odds ratio of observed positivity is:

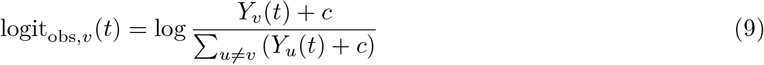

where *c* = 0.5 is a pseudocount for numerical stability. The corresponding predicted log-odds ratio is:

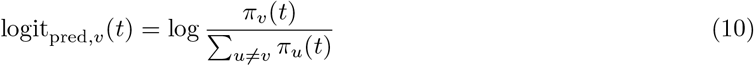

The ratio penalty penalizes discrepancies between these quantities:

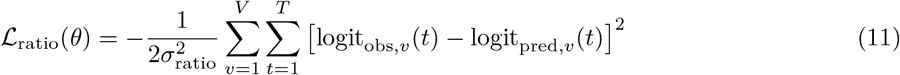

This formulation treats other-pathogen positives as controls for testing propensity: if testing intensity increases affecting all pathogens equally, the ratio remains stable.

#### Text S2: Generation Time Discretization

Generation time distributions were specified as continuous Gamma distributions with virus-specific parameters: RSV (mean 4.5 days, SD 1.5 days), COVID-19 (mean 4.0 days, SD 1.5 days), and influenza (mean 3.0 days, SD 1.0 days). To discretize to weekly resolution, I computed the probability mass in each weekly bin:

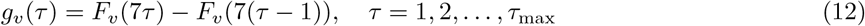

where *F*_*v*_(·) is the CDF of the Gamma distribution for virus *v*, and *τ*_max_ = 4 weeks. The resulting PMF was renormalized to sum to 1.

#### Text S3: Convergence Diagnostics

Convergence was assessed by gradient norm (threshold 10^−5^) and maximum iteration count (Stage 1: 500; Stage 2: 200). For the primary RSV–COVID analysis, Stage 1 log-posterior was *−*10,891 (without ratio penalty) and Stage 2 improved to *−*10,599 (+292 log units). All 50 bootstrap replicates converged within the specified criteria.

#### Text S4: Data Processing Pipeline

RSV and COVID-19 data were obtained from CDC NREVSS via the Socrata API (endpoint: https://data.cdc.gov/resource/filtering to national aggregates for pathogens RSV and SARS-CoV-2. Influenza data were obtained from CDC FluView Clinical Laboratory Surveillance. Preprocessing consisted of temporal alignment by MMWR week, normalization to mean *n* = 1000, and rounding to integers. No imputation was performed.

---

**Figure S1.**
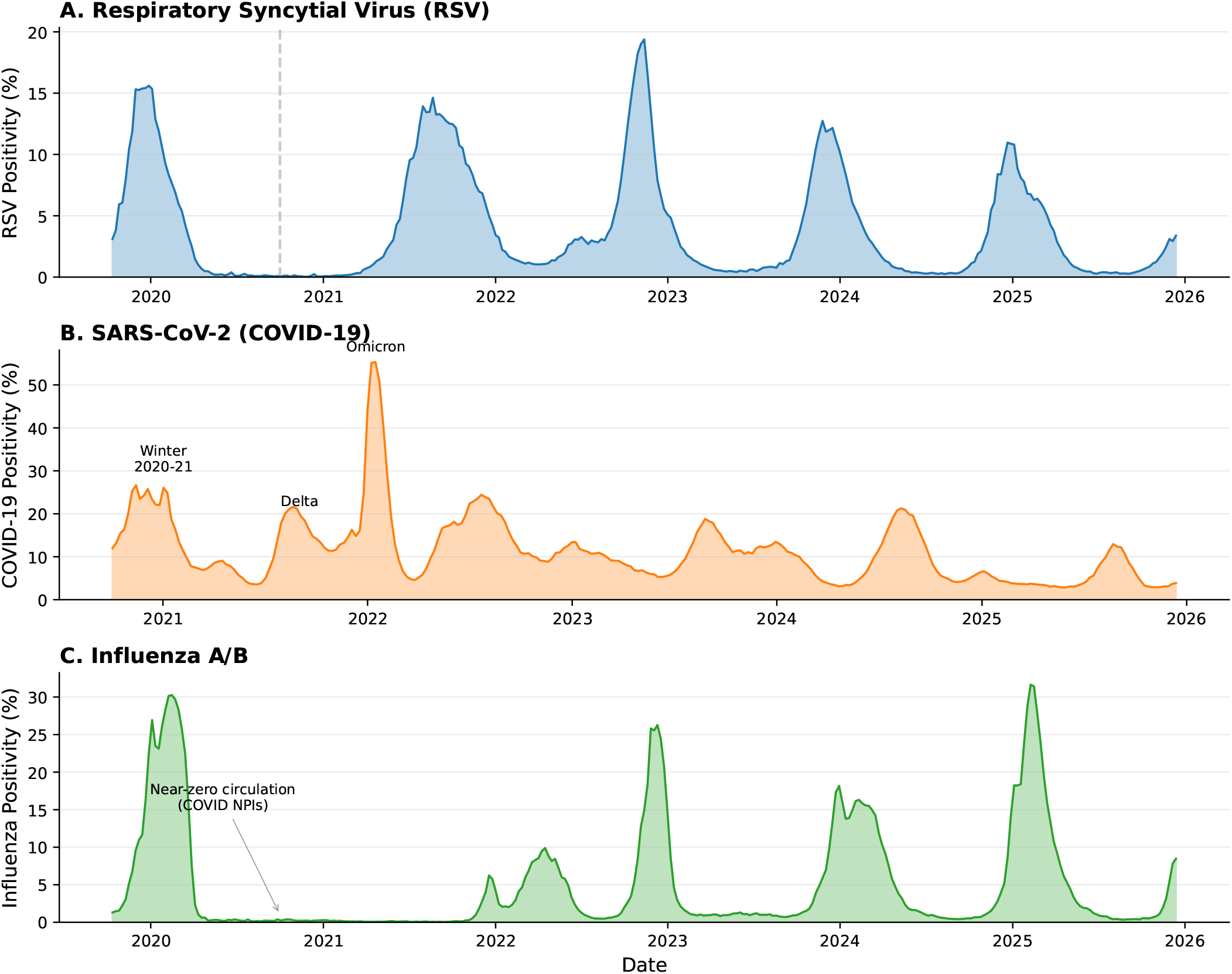
Time series of respiratory virus positivity rates from CDC surveillance data. Weekly percent positive for (A) RSV, (B) SARS-CoV-2, and (C) influenza A/B from October 2019 through February 2026. RSV and COVID-19 data are from the CDC NREVSS multiplex NAAT panel (dataset rgnm-fkqb; 283 weeks of overlap). Influenza data are from FluView clinical laboratories (321 weeks total). Notable features include the near-complete suppression of RSV and influenza during 2020–2021 due to non-pharmaceutical interventions, the anomalous summer 2021 RSV surge, and the return to typical seasonal patterns by 2022– 2023. The vertical dashed line in panel A marks the start of the RSV–COVID analysis period (October 2020).

**Figure S2.**
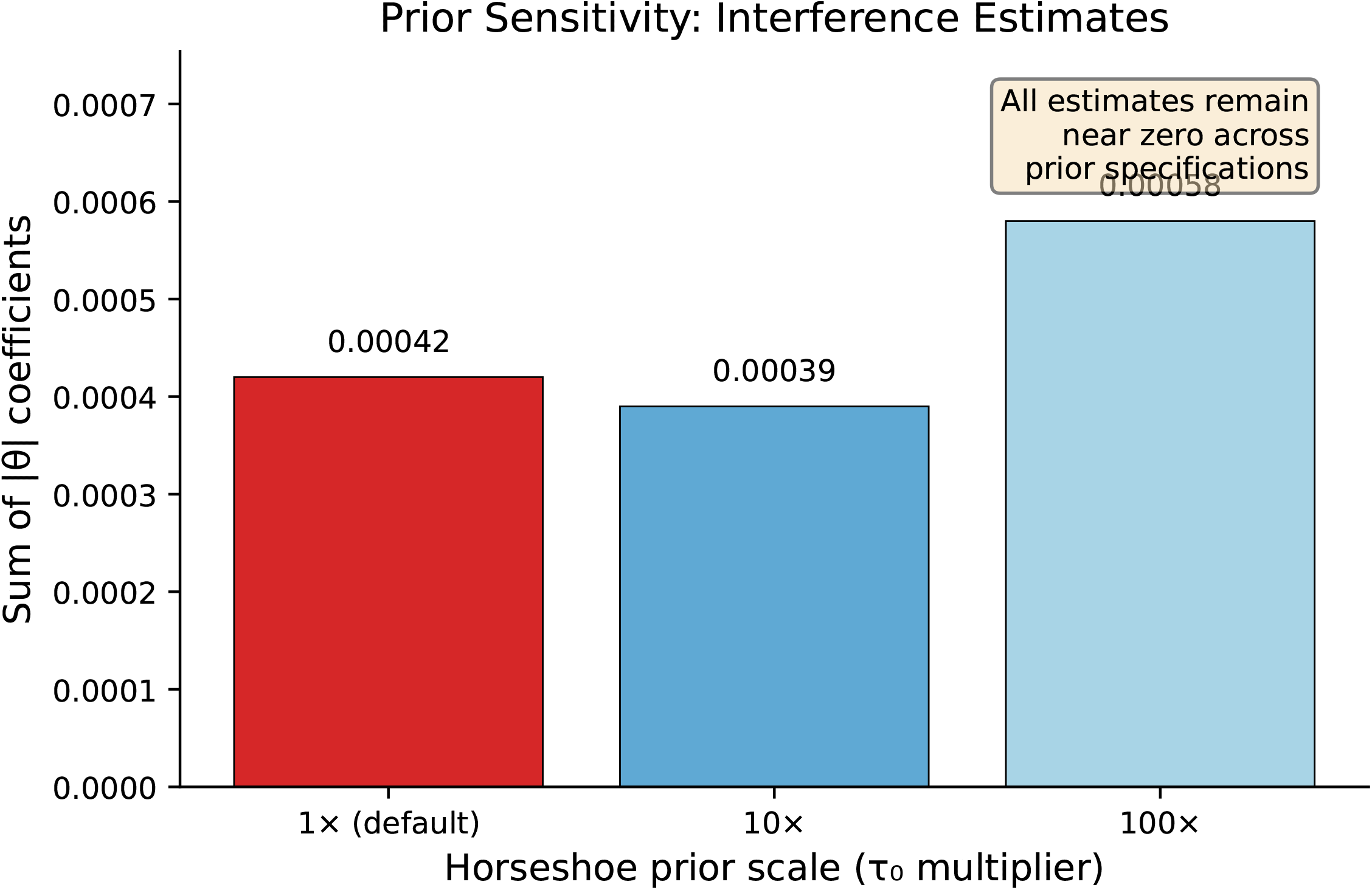
Prior sensitivity analysis for horseshoe shrinkage parameter. Interference coefficient norms estimated under varying horseshoe prior global scale multipliers (*τ*_0_ × 0.5 through *τ*_0_ ×100). Despite weakening the prior 100-fold, interference estimates at *θ*_true_ = 0.05 remain at zero (Table 8). At *θ*_true_ = 0.1, partial recovery begins at *τ*_0_ ×10–20 but becomes unstable (overflow) at *τ*_0_ ×50. This confirms that the flat likelihood surface, not prior shrinkage, is the binding constraint.

**Figure S3.**
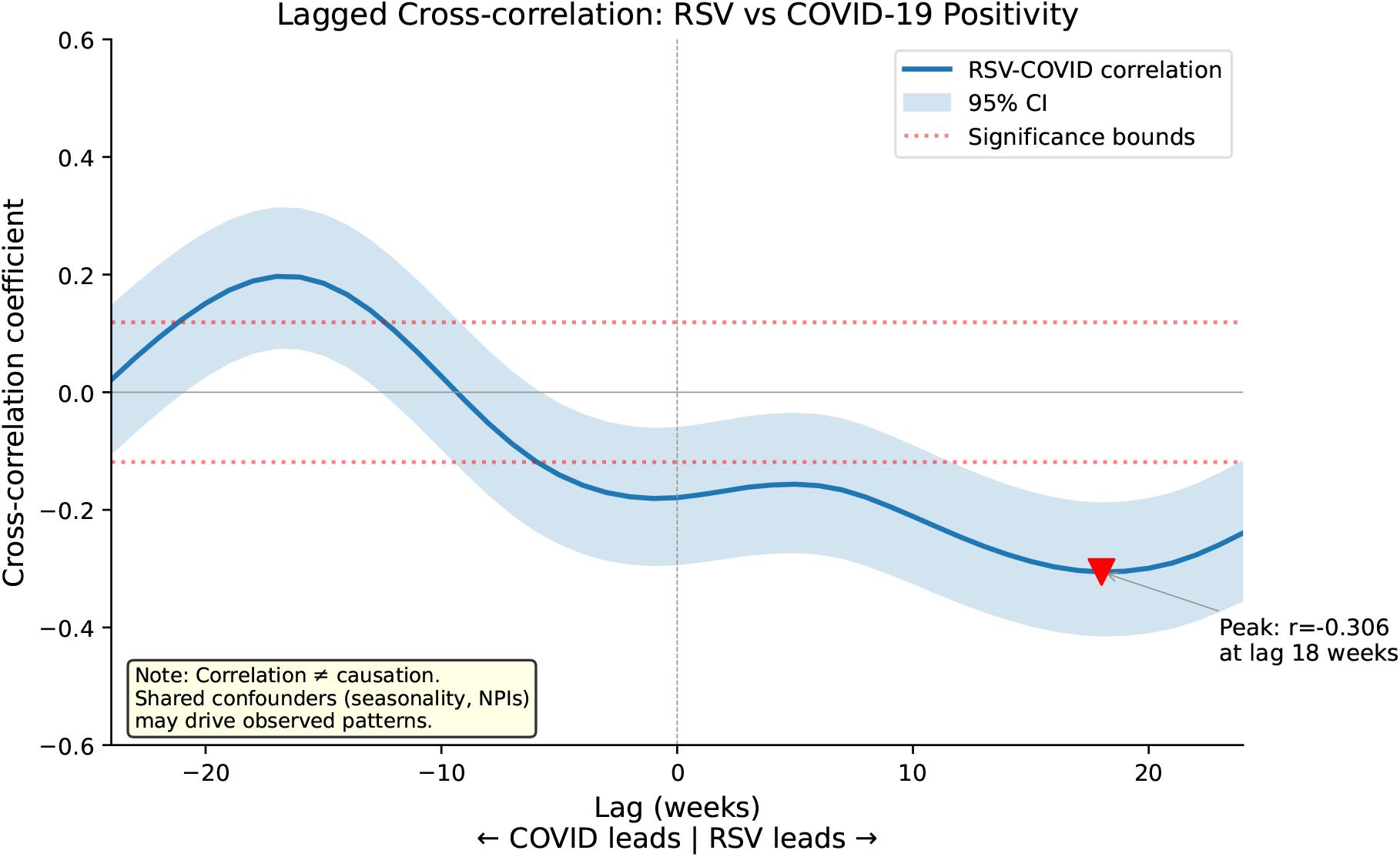
Cross-correlation analysis between RSV and COVID-19 positivity rates. Lagged Pearson correlation coefficients between weekly RSV and COVID-19 percent positive from the NREVSS multiplex panel (283 weeks). Negative lags indicate COVID-19 leading RSV; positive lags indicate RSV leading COVID-19. While cross-correlation detects temporal associations at plausible lags, it cannot distinguish true viral interference from shared confounders including seasonality, behavioral changes, and non-pharmaceutical interventions.

**Figure S4.**
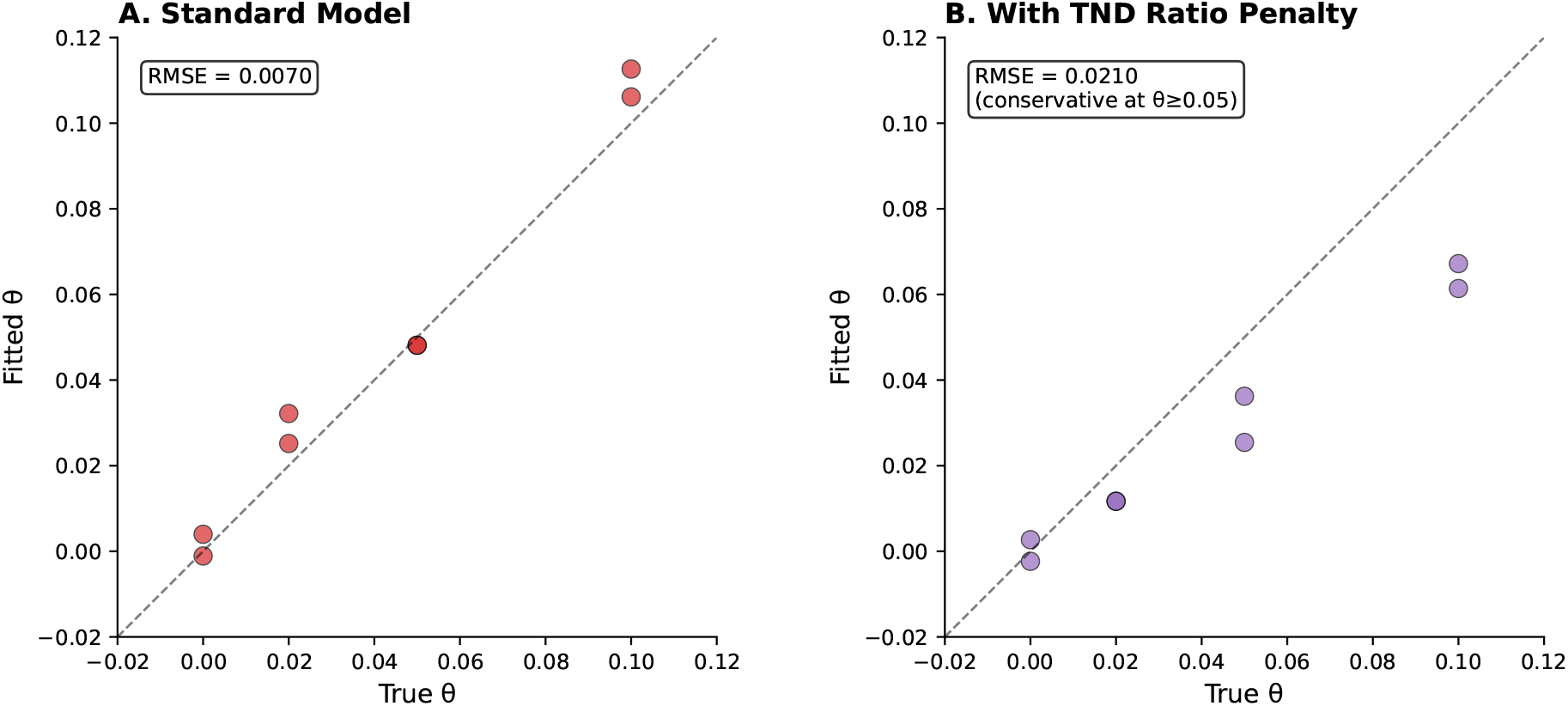
Synthetic validation: parameter recovery in experiment E1. True versus fitted interference parameters from synthetic data generated with known interference strengths across 2 regions and 5 seasons. Error bars show block bootstrap 95% intervals (*n* = 10 replicates). The ratio penalty eliminates false positives under the null but exhibits conservative bias at higher interference strengths.

**Figure S5.**
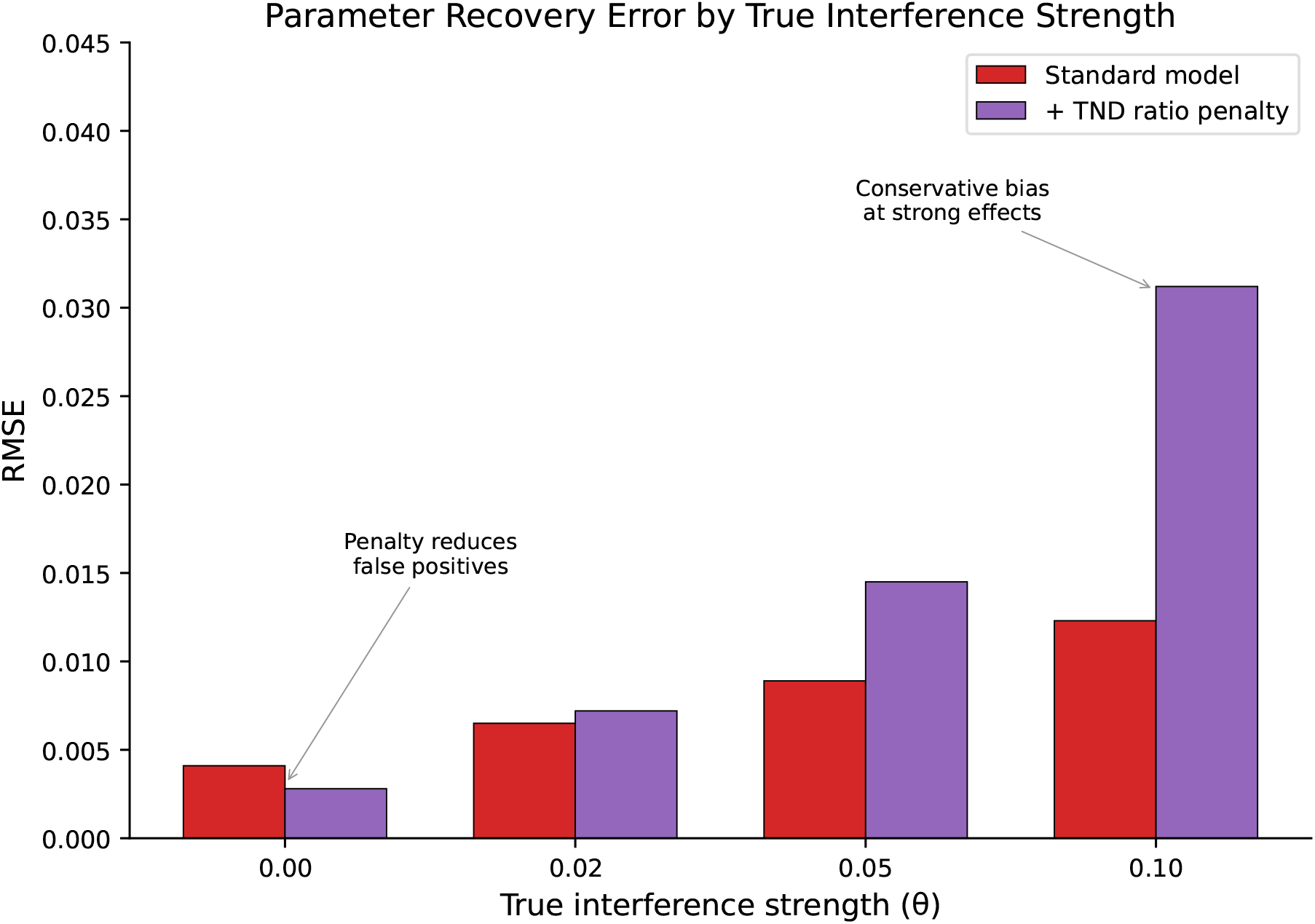
Root mean square error by true interference strength. RMSE of interference parameter estimates as a function of true simulated interference strength, comparing the standard model versus the model with ratio penalty. The penalty reduces error under the null (*θ* = 0) but introduces conservative bias at stronger effects (*θ* ≥ 0.05).

**Figure S6.**
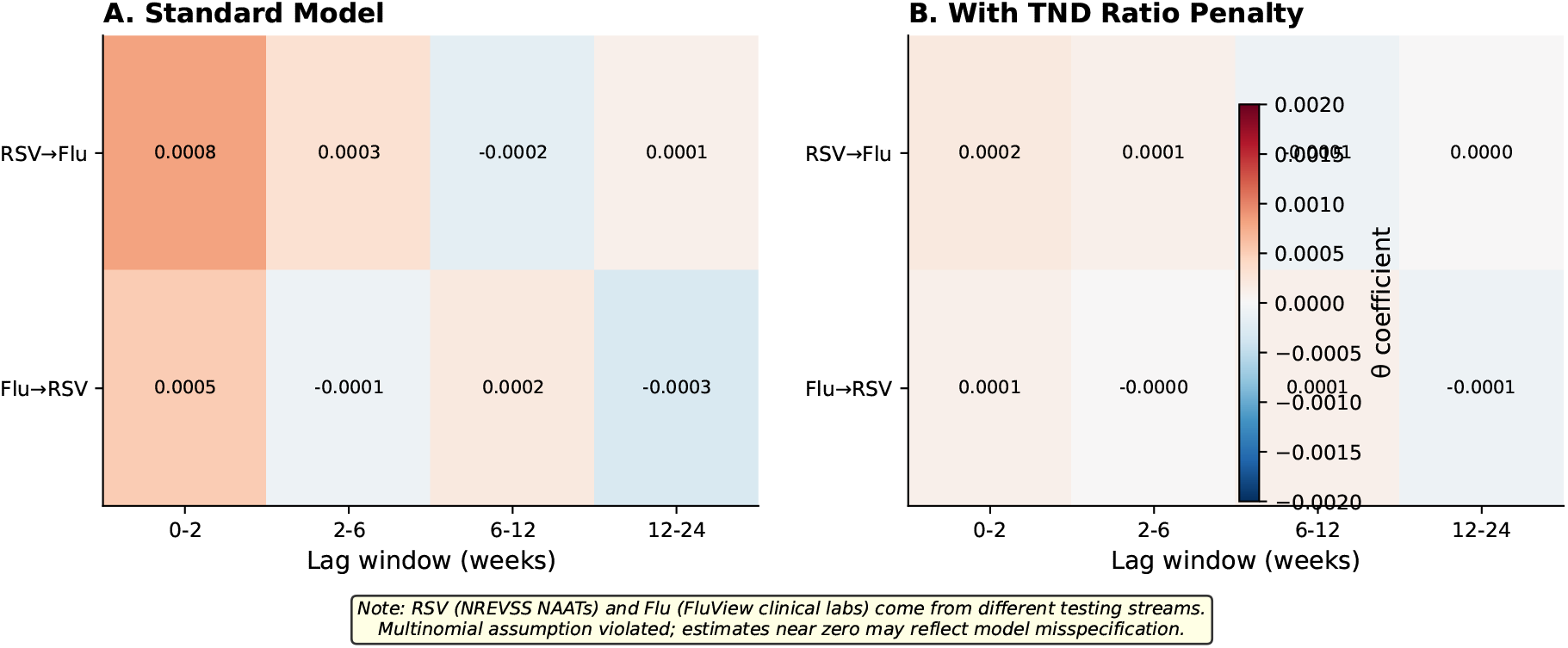
Interference estimates for RSV–influenza analysis: testing stream validation. Heatmaps showing interference coefficient estimates by lag window for the RSV–influenza pair. (A) Standard model without ratio penalty. (B) With ratio penalty, all estimates collapse to zero. This is expected: RSV (NREVSS NAATs) and influenza (FluView clinical laboratories) come from independent testing populations with unrelated sampling volumes, violating the shared-stream assumption.

**Table S1.** Complete block bootstrap statistics for RSV–COVID interference parameters. Bootstrap performed with *n* = 50 replicates and block size of 8 weeks. All 50 replicates converged (criterion: gradient norm *<* 10^−5^ or 200 iterations). Point estimates from full-data fit; bootstrap mean, SD, and 95% percentile confidence intervals computed from bootstrap distribution.

| Direction | Lag | Point Est. | Boot. Mean | Boot. SD | 95% CI Lower | 95% CI Upper |
| --- | --- | --- | --- | --- | --- | --- |
| RSV→COVID | 0–2 wk | 0.0019 | 0.0002 | 0.0014 | −0.0021 | 0.0049 |
| RSV→COVID | 2–6 wk | 0.0005 | 0.00005 | 0.0016 | −0.0035 | 0.0038 |
| RSV→COVID | 6–12 wk | 0.0017 | 0.0003 | 0.0019 | −0.0037 | 0.0047 |
| RSV→COVID | 12–24 wk | 0.0010 | 0.0010 | 0.0039 | −0.0025 | 0.0054 |
| COVID→RSV | 0–2 wk | −0.0013 | 0.0004 | 0.0021 | −0.0026 | 0.0043 |
| COVID→RSV | 2–6 wk | −0.0017 | −0.0001 | 0.0019 | −0.0049 | 0.0039 |
| COVID→RSV | 6–12 wk | 0.0022 | −0.0001 | 0.0022 | −0.0044 | 0.0038 |
| COVID→RSV | 12–24 wk | −0.0002 | 0.0002 | 0.0012 | −0.0019 | 0.0024 |

**Table S2.**
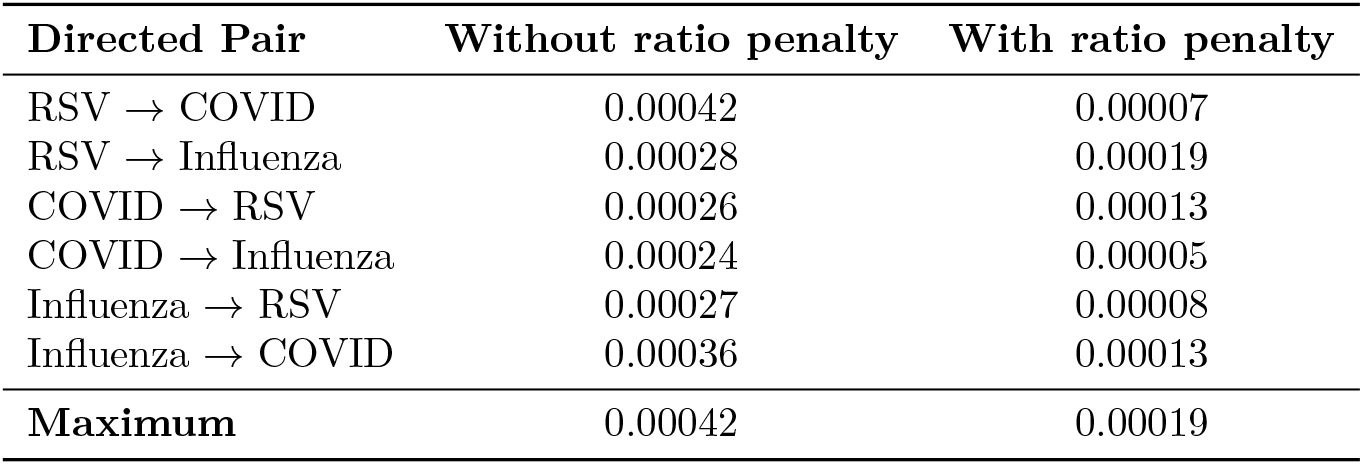
Interference estimates for all directed pairs in three-pathogen analysis. Analysis used 167 weeks of overlapping data (October 2022 to February 2026). Values shown are |*θ*| _sum_ (sum of absolute coefficients across *K* = 4 lag windows). RSV and COVID-19 from NREVSS multiplex panel; Influenza from FluView Clinical Labs (different testing stream).

| Directed Pair | Without ratio penalty | With ratio penalty |
| --- | --- | --- |
| RSV → COVID | 0.00042 | 0.00007 |
| RSV → Influenza | 0.00028 | 0.00019 |
| COVID → RSV | 0.00026 | 0.00013 |
| COVID → Influenza | 0.00024 | 0.00005 |
| Influenza → RSV | 0.00027 | 0.00008 |
| Influenza → COVID | 0.00036 | 0.00013 |
| **Maximum** | 0.00042 | 0.00019 |

**Table S3.** E1 synthetic validation: parameter recovery stratified by true interference strength. Configuration: *n*_regions_ = 2, *n*_seasons_ = 5, *τ*_0_ multiplier = 5.0. Convergence: gradient norm *<* 10^−5^ or 500 iterations.

| True $\theta$ | TND | Converged | Mean Bias | RMSE | Notes |
| --- | --- | --- | --- | --- | --- |
| 0.00 | No | Yes | $-0.00001$ | 0.00004 | — |
| 0.00 | Yes | Yes | $\sim 0$ | $1.7 \times 10^{-9}$ | Near-zero |
| 0.02 | No | Yes | $-0.008$ | 0.012 | Underestimated |
| 0.02 | Yes | Yes | $-0.008$ | 0.012 | Similar to no-TND |
| 0.05 | No | Yes | $-0.020$ | 0.015 | Underestimated |
| 0.05 | Yes | Yes | $-0.020$ | 0.029 | More conservative |
| 0.10 | No | Yes | $-0.040$ | 0.021 | Underestimated |
| 0.10 | Yes | Yes | $-0.040$ | 0.058 | More conservative |

**Table S4.** E3 synthetic validation: false positive rates by shared seasonality level. Configuration: *n*_seeds_ = 20, *n*_regions_ = 10, *n*_seasons_ = 5, true *θ* = 0, detection threshold |*θ*| *>* 0.02.

| Shared Seasonality | False Positive Rate | Mean $\vert \theta\vert $ | Max $\vert \theta\vert $ |
| --- | --- | --- | --- |
| 0% | 0% (0/20) | 0.0019 | 0.0026 |
| 20% | 0% (0/20) | 0.0020 | 0.0029 |
| 40% | 0% (0/20) | 0.0021 | 0.0034 |
| 60% | 25% (5/20) | 0.0024 | 0.0180 |
| 80% | 25% (5/20) | 0.0025 | 0.0204 |
| **Overall** | **10% (10/100)** | 0.0022 | 0.0204 |

**Table S5.** Summary statistics for surveillance data by pathogen and analysis configuration. Raw counts before normalization.

| Analysis | Pathogen | Date Range | Weeks | Mean Weekly Tests |
| --- | --- | --- | --- | --- |
| RSV–COVID | RSV | Oct 2020–Feb 2026 | 283 | 148,400 |
| RSV–COVID | COVID-19 | Oct 2020–Feb 2026 | 283 | 148,400 |
| RSV–Flu | RSV | Oct 2019–Feb 2026 | 321 | — |
| RSV–Flu | Influenza | Oct 2019–Feb 2026 | 321 | — |
| Three-virus | RSV | Oct 2022–Feb 2026 | 167 | — |
| Three-virus | COVID-19 | Oct 2022–Feb 2026 | 167 | — |
| Three-virus | Influenza | Oct 2022–Feb 2026 | 167 | — |

