## Supplementary figures and images for "Apparent RSV–COVID interference is not robust to adjustment for shared testing propensity"

### Supplemental Figure 1

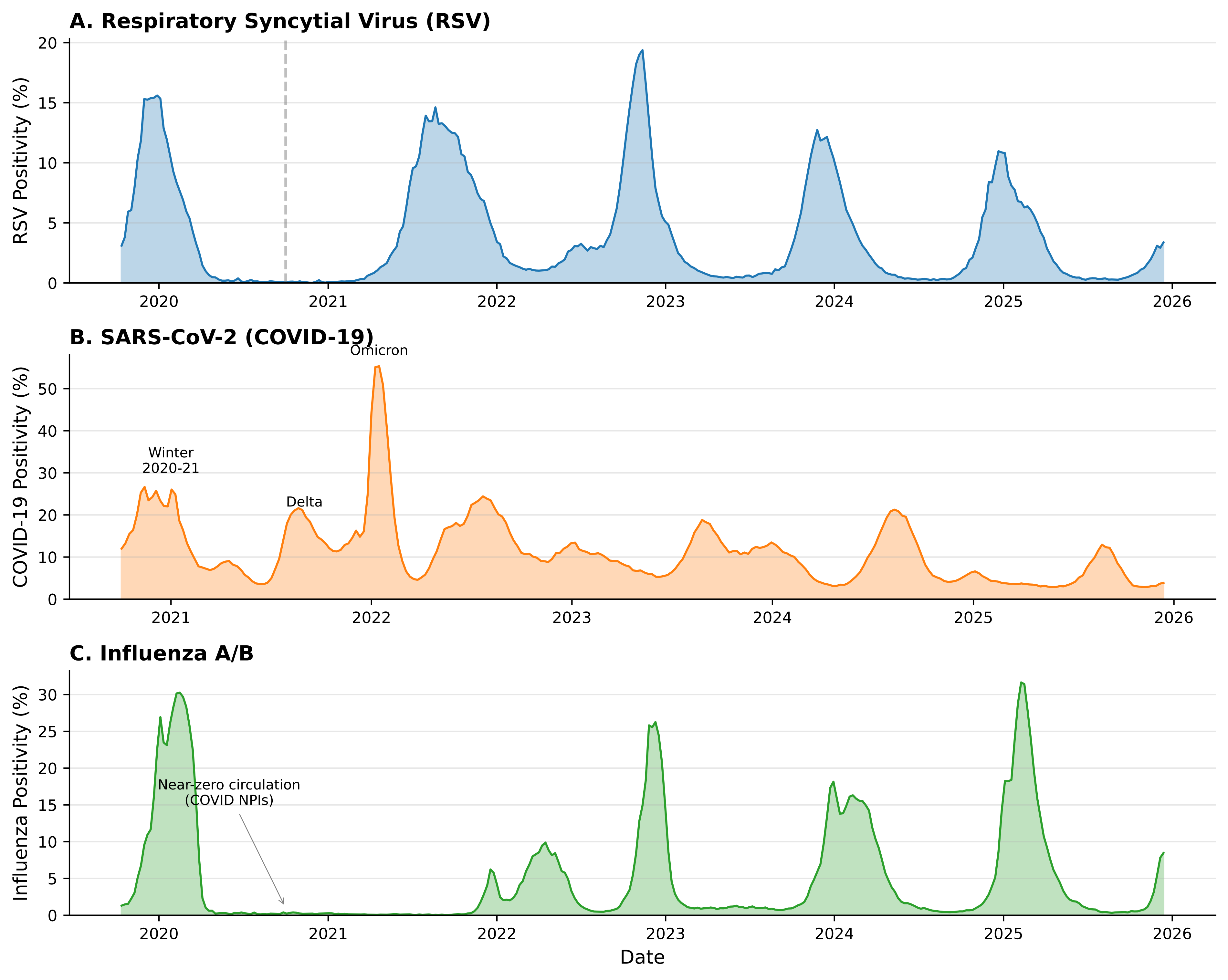

### Supplemental Figure 2

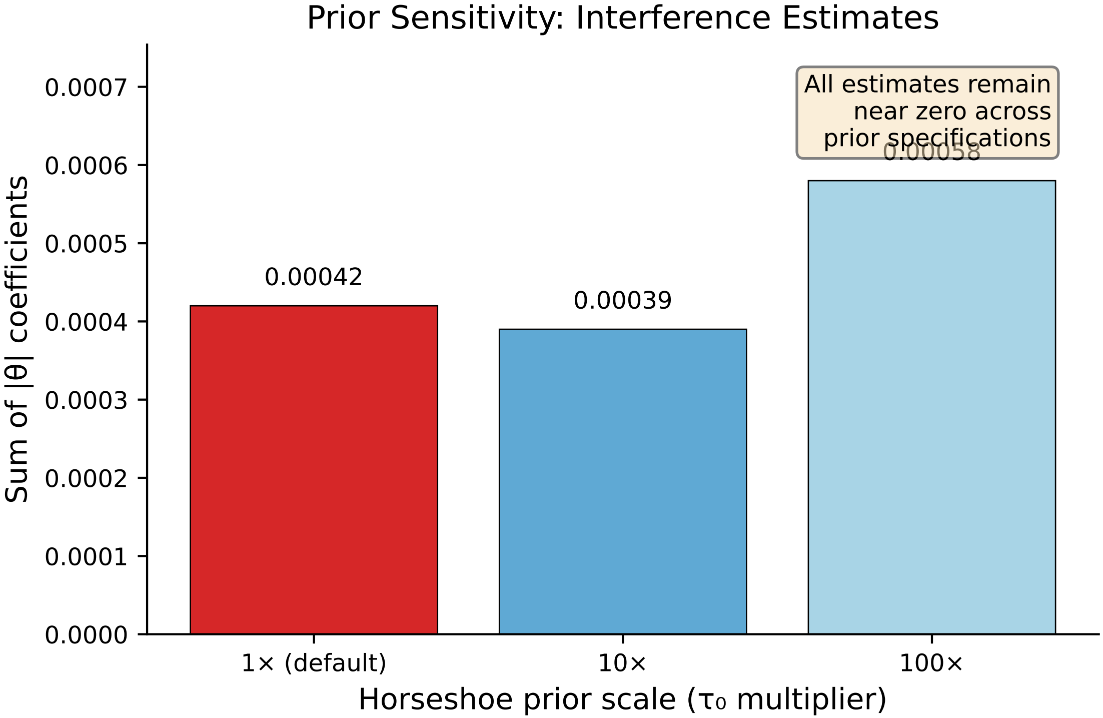

### Supplemental Figure 3

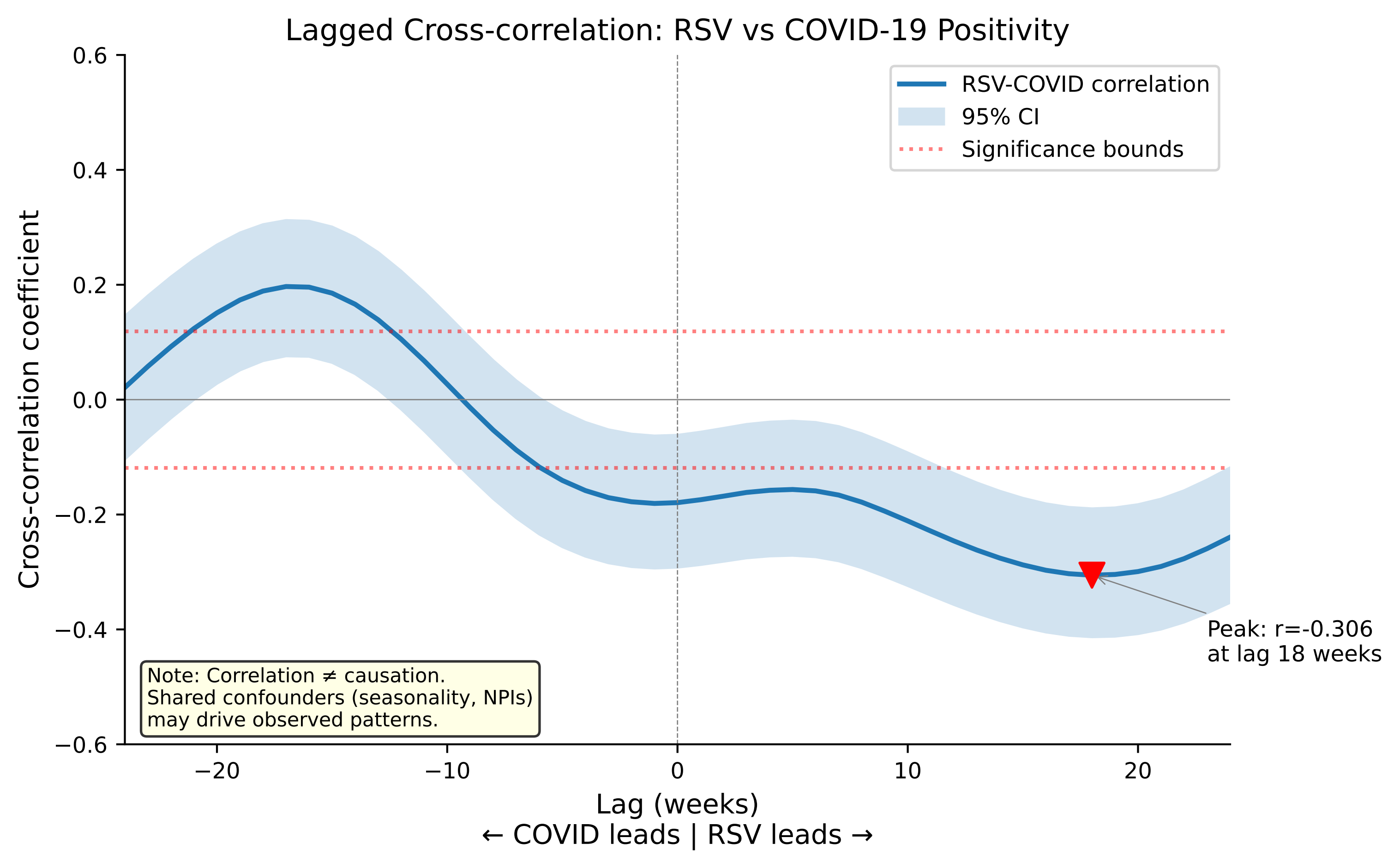

### Supplemental Figure 4

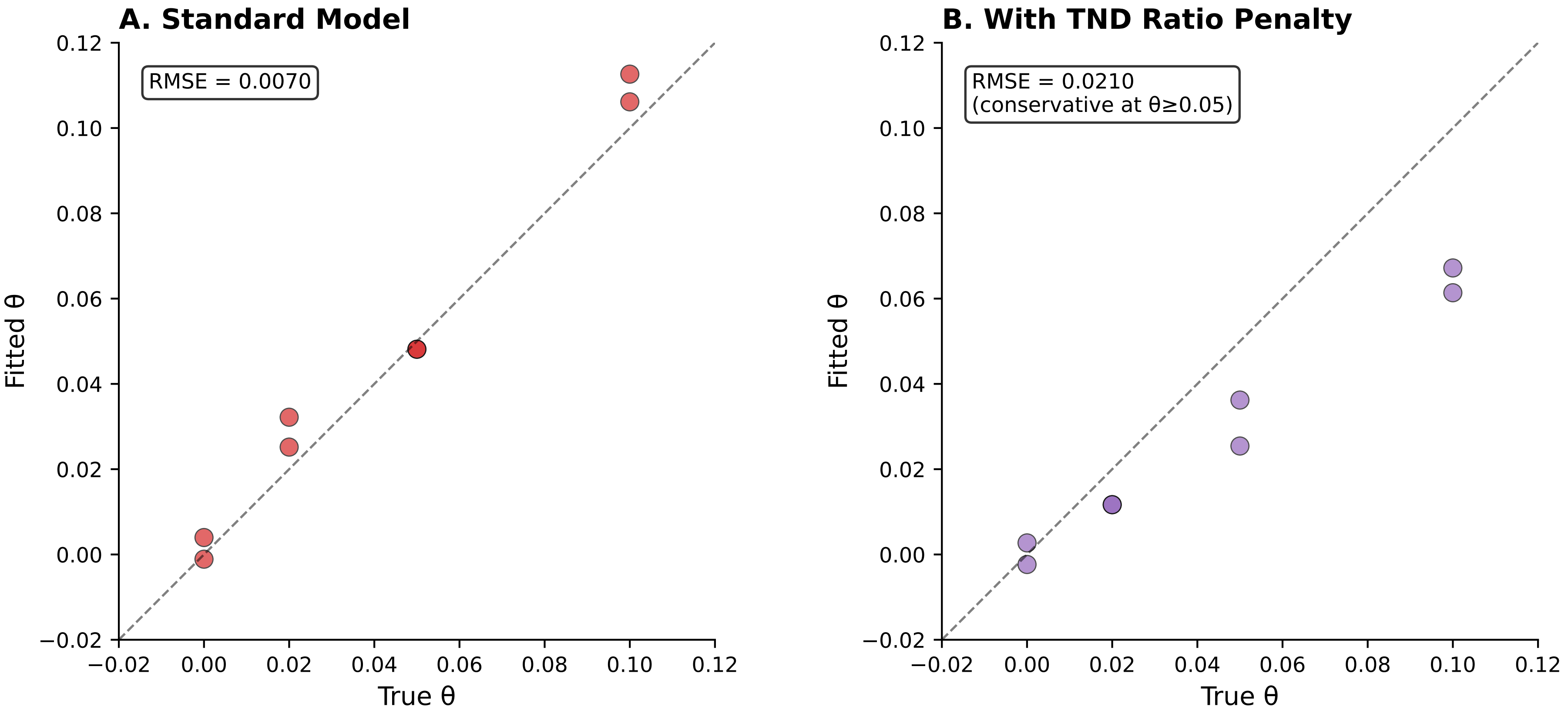

### Supplemental Figure 5

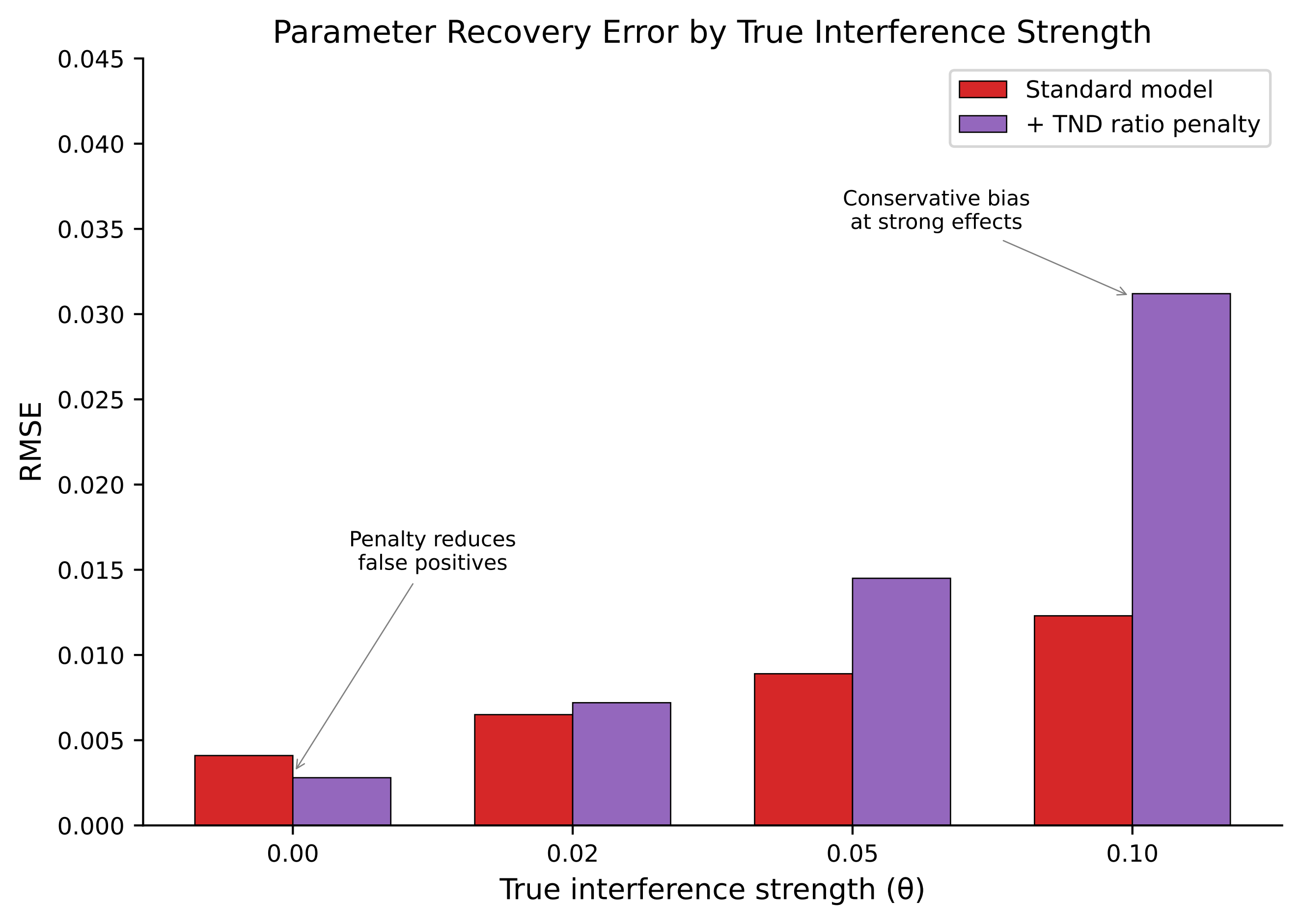

### Supplemental Figure 6

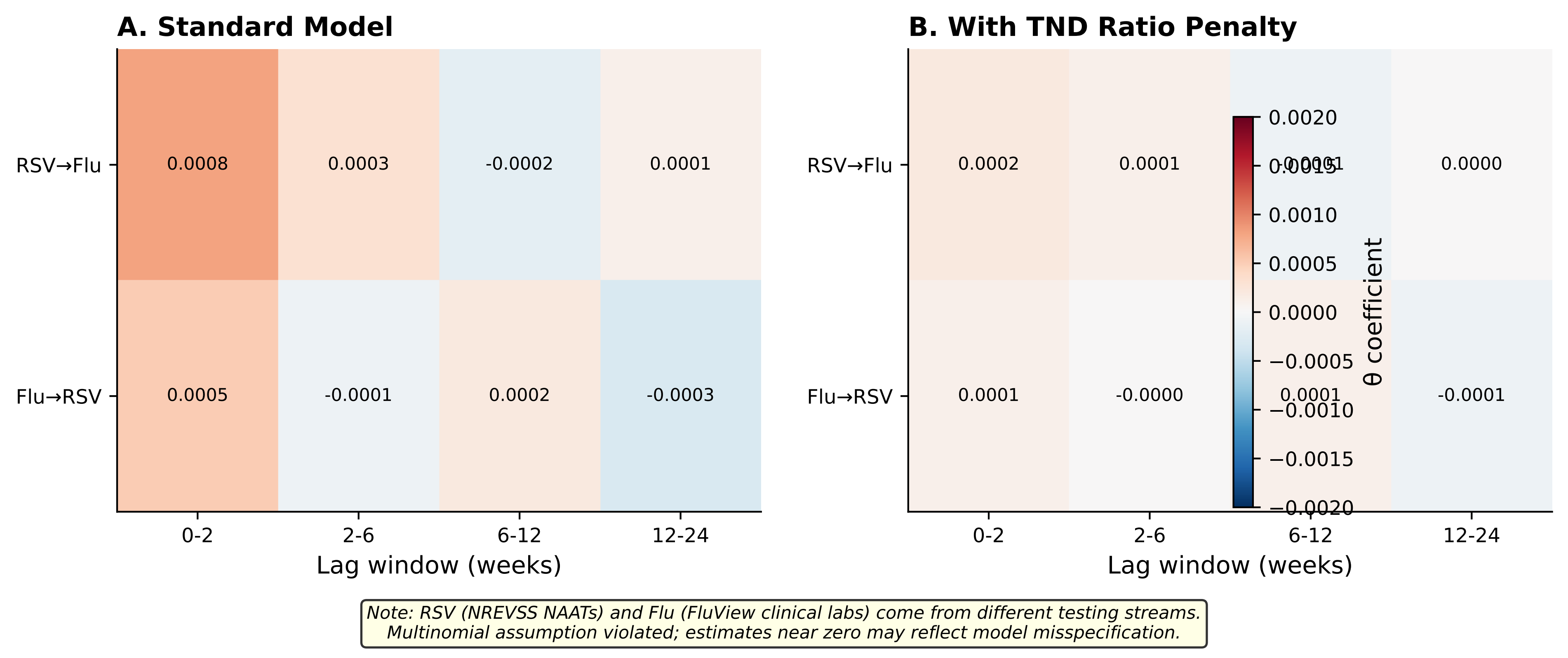
